# Inequalities and practice-level predictors of cancer diagnostic activity: longitudinal study in England

**DOI:** 10.64898/2026.09.04.26362238

**Authors:** Alfariany Fatimah, Samuel WD Merriel, Emma Whitfield, Becky White, Georgios Lyratzopoulos, Igor Francetic

## Abstract

**Background:** Early cancer detection is aided by timely GP referrals. However, GP practices face pressures, particularly in deprived areas, which may be associated with diagnostic inequalities.

**Aim:** To examine whether practice-level indicators of access, workforce, and population income deprivation were associated with diagnostic care indicators.

**Design and setting:** Retrospective longitudinal study of 6,194 practices between 2015-2019 and 2022-2023.

**Method:** Panel fixed-effects regression examined USC referral rates, USC PPV, USC sensitivity, and emergency presentation rates. Predictors were patient-reported access, GP workforce, and practice population income deprivation.

**Results:** Per one SD increase within practices, (i) USC referral rates were lower with better appointment-making experience (-1.4%) and older average GP age (-3.5%), and higher with female GP share (+1.3%); GP FTE showed a small positive association (+0.7% to +0.8%). (ii) USC PPV increased with older average GP age (+2.2%). (iii) USC sensitivity was lower with older average GP age (-1.1%); emergency presentations showed limited association with access or workforce factors. Practices had higher emergency presentation rates when serving the most deprived rather than least deprived populations (+13.7%). There was little evidence that deprivation modified access and workforce associations. Average GP age was an exception: USC referral associations weakened from Q2 to Q5, USC PPV peaked at Q3, and USC sensitivity was significant in the most deprived quintile.

**Conclusion:** Practice-level access and workforce factors were associated with USC referral activity. Persistent deprivation gaps in emergency presentation suggest value in monitoring practice-level USC referrals alongside their PPV and sensitivity, and practice-level burden of emergency presentations.

**How this fits in:** USC referral rates vary widely between general practices, and previous studies link this variation to GP age, staffing, access, practice size, QOF achievement, and deprivation (1–3). Most evidence is cross-sectional, so it is unclear whether these factors explain stable differences between practices or changes within the same practice over time. We showed that access and workforce factors were associated mainly with referral volume, while deprivation was more strongly linked to practice-level burden of emergency presentation than to practice-level referral activity. Persistent deprivation gaps in emergency presentations, despite narrowing referral-rate gaps, suggest that equity efforts should look beyond referral volume alone.

## Introduction

Approximately 7-in-10 patients with cancer in England are diagnosed following contact with NHS primary care. Their route to diagnosis can be via the urgent suspected cancer (USC) referral pathways, routine outpatient referrals, and emergency presentations (4). The use of USC referrals has been increasing over the last decade, as has the proportion of cancers detected via USC referral (5). This has been accompanied by a decrease in the proportion of cancers detected via emergency presentation (6). Higher rates of USC referrals at GP practice level are associated with earlier stage at diagnosis and increased 5-year survival (7). These trends are consistent with improvements in cancer diagnosis through NHS primary care.

USC referral use varies widely between practices and is linked to practice-level characteristics. Practices with more nurses (8), younger GPs (2), better provider-patient communication (3), and training practices (2) were associated with higher urgent cancer referral rates in previous cross-sectional studies. Practices with larger patient lists, younger GPs, higher *Quality and Outcomes Framework* (QOF) scores, and a higher proportion of older patients (>65 years) have been found to achieve higher sensitivity of USC referrals (1). Conversely, practices with higher patient-to-GP ratios, older GP workforces, greater deprivation, and higher proportions of non-white ethnicity tend to have lower USC sensitivity (1). Being based on cross-sectional designs, these findings are prone to bias.

We used a longitudinal approach and examined how within-practice changes over time in patient-reported phone access, appointment-making experience, and workforce composition were associated with four indicators of cancer diagnostic activity. Alongside USC referral rates, we examined positive predictive value (‘conversion rate’, USC PPV), sensitivity (‘detection rate’, USC sensitivity), and emergency presentation rates as a proxy for late diagnosis. We also examined whether association between these predictors and our outcomes varied by practice-population income deprivation levels.

## Methods

### Data

We conducted a longitudinal analysis of publicly available practice-level data from general practices, covering 2015–2019 and 2022–2023. This was the longest time window available after linking yearly data on all variables. We excluded 2020 and 2021 to avoid pandemic-related disruptions. We included 6,194 of 6,219 practices; 25 were excluded because outcome or predictor data were unavailable. Included practices were active practices in the National General Practice Profiles (Fingertips), defined as having a registered list size of at least 750 patients. Within these included practices, we excluded practice-year observations with list size above the pooled 99th percentile and average GP age below the pooled 1st percentile, removing observations rather than whole practices. The panel was near-balanced: 93% of practices were observed in all analysis years; the rest reflected openings, closures, or mergers.

Table 1 lists the outcome variables, predictors, control variables, and data sources. **Supplementary Material Section A and Table S.1** describe data processing and variable construction in more detail.

**Table 1.**
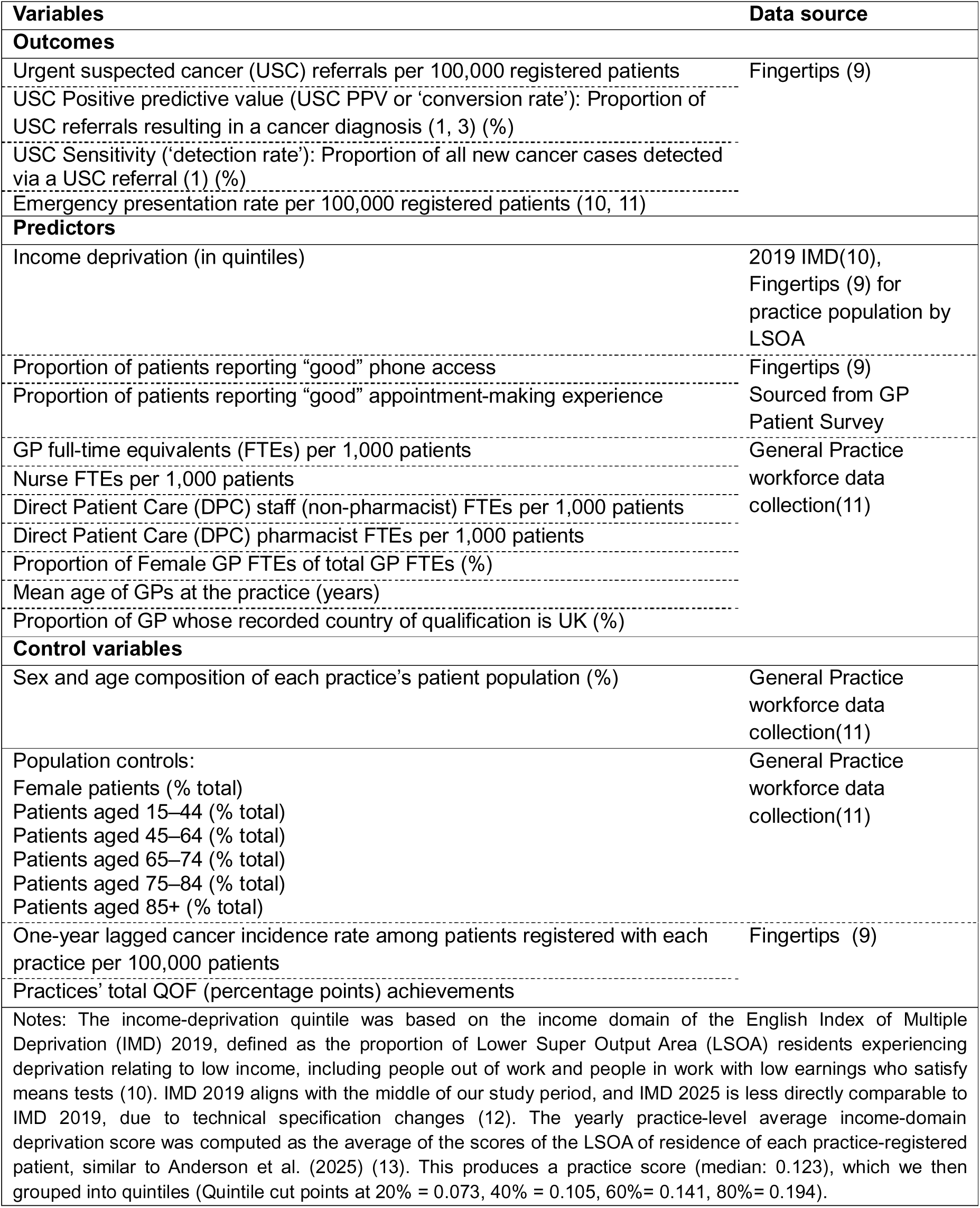
Outcomes and predictors.

### Statistical approach

We examined USC referral rates, USC PPV, USC sensitivity, and emergency presentation rates using regression models estimated with a fixed effects (within) estimator, hence exploiting variation within-practices over time. Across all regression models, our key predictors were access indicators (phone access and appointment-making experience) and workforce indicators (proportion female GP, average GP age, GP UK medical qualification, and GP, nurse, DPC non-pharmacist, and DPC pharmacist FTEs per 1,000 patients). The key predictors were chosen because access may be related to whether people seek care, while workforce composition may be linked to referral decisions. All models include the following control variables: lagged crude cancer incidence, QOF points, and practice population sex and age composition, and calendar-year fixed effects. Given our estimator of choice, all models include GP practice fixed effects.

For each outcome, we estimated two regression models. In Model 1, we included practice population income deprivation additively, as a predictor. In Model 2, we interacted practice population income deprivation with all other predictors and control variables, allowing estimated associations of key predictors to vary across deprivation levels through interaction terms. **Supplementary material section B** provides details on the econometric modelling. All results for Model 2 are reported as average marginal effects, overall or by deprivation level. To improve interpretability, all associations were rescaled to percentage changes in the outcome for a one-standard-deviation increase in each predictor, relative to the sample mean. This enabled comparisons across predictors measured on different scales.

Statistical inference is based on robust standard errors clustered at GP practice level. All analyses were conducted using the statistical software Stata 19. We adjusted for multiple hypothesis testing using Šidák correction in the main models and robustness analyses.

### Robustness analyses

We assessed robustness using subgroup analyses (rurality, ethnic composition, and common cancer sites: breast, lower GI, lung, and skin) and sensitivity to some design choices (splitting pre- and post-COVID-19, accounting for screening uptake, practice size, using deciles instead of quintiles, and adding USC referral volume as a control in PPV models). The latter analysis tested whether the estimated association reflected a relationship between predictors and diagnostic accuracy or a mechanical effect driven by referral volume in the denominator. Full definitions and rationale for all analyses are provided in **Supplementary Material Section E**.

## Results

Table 2 reports descriptive statistics for the full sample of 39,421 practice-year observations across 6,194 practices, with an average of 6.36 yearly observations per practice. Within-practice standard deviations were generally substantial relative to between-practice standard deviations. USC referral rates increased, while USC PPV and access measures declined between 2015 and 2023. USC sensitivity and emergency presentation rates remained broadly stable. Practices serving more deprived populations had higher USC referral rates but lower USC PPV and USC sensitivity than practices serving less deprived populations, with the gap shifting over time. (**Supplementary Material Section C, Figures S1 and S2).**

**Table 2.**
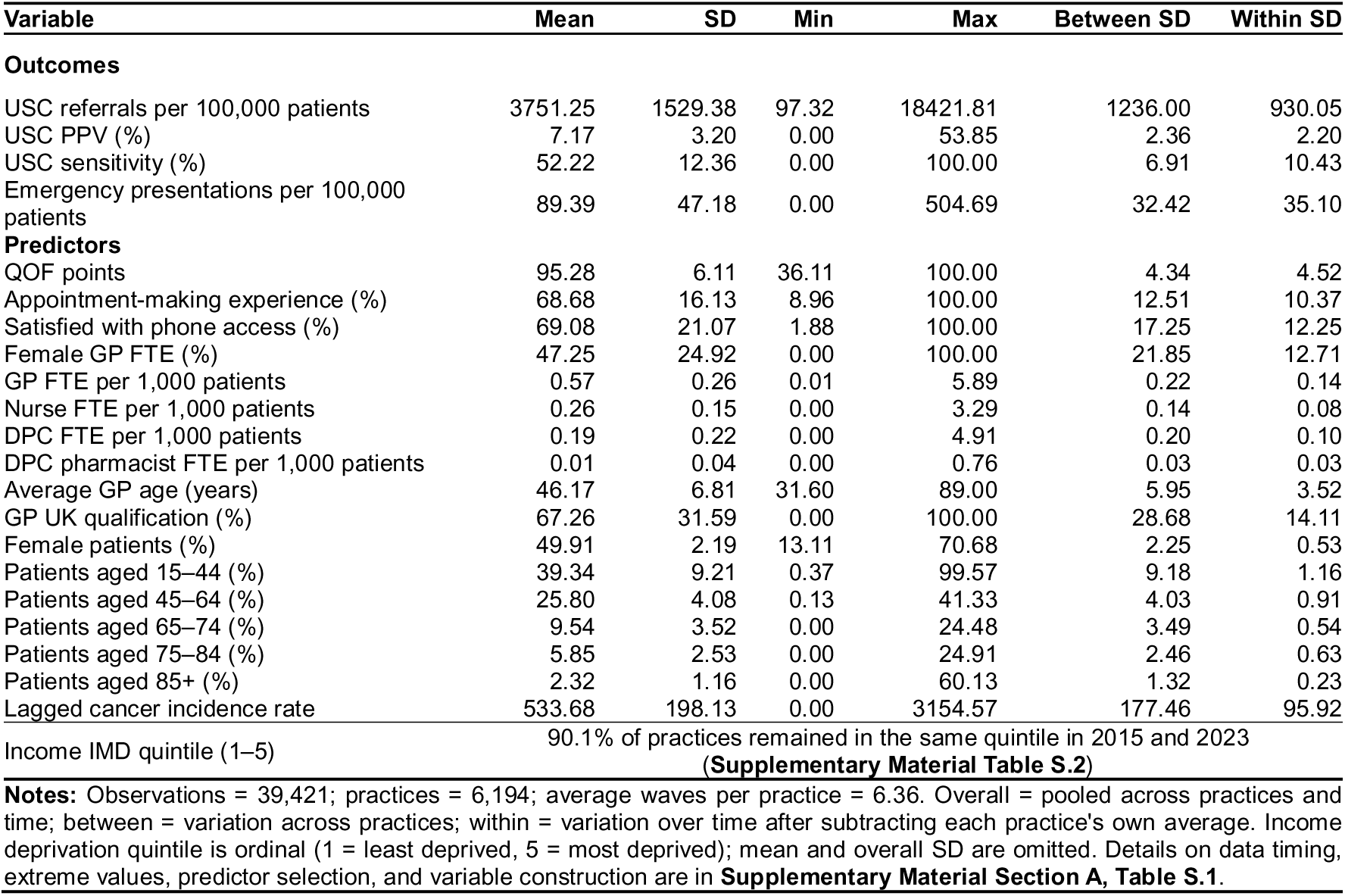
Summary statistics of outcomes and predictors.

Table 3 reports results from Model 1 compared with Model 2, the deprivation-interaction fixed-effects model. Focusing on Model 2, USC referral rates were negatively associated with appointment-making experience (-1.4%) and phone access (-0.8%); the latter did not remain significant after Šidák correction. By contrast, USC referral rates were positively associated with within-practice increases in GP FTEs (+0.7%) and female GP share (+1.3%). Older average GP age was associated with fewer USC referrals (-3.5%); the corresponding Model 1 estimate was -3.2%. For USC PPV, average GP age was the only access or workforce predictor that survived multiple-testing correction in Model 2 (+2.2%). For USC sensitivity, older average GP age was also negatively associated with USC sensitivity in Model 2 (-1.1%), and this association survived Šidák correction. Appointment-making experience was associated with lower emergency presentation rates in Model 2 (-1.3%), although this did not survive Šidák correction.

**Table 3.** Key predictors: percentage changes in cancer diagnostic outcomes.

| Predictor | USC Referral |  | USC PPV |  | USC Sensitivity |  | Emergency Presentation |  |
| --- | --- | --- | --- | --- | --- | --- | --- | --- |
|  | Model 1 (Non-int.) | Model 2 (Int. × IMD) | Model 1 (Non-int.) | Model 2 (Int. × IMD) | Model 1 (Non-int.) | Model 2 (Int. × IMD) | Model 1 (Non-int.) | Model 2 (Int. × IMD) |
| Mean outcome | 3,751 |  | 7.17 |  | 52.22 |  | 89.4 |  |
| Units | Per 100,000 |  | % |  | % |  | Per 100,000 |  |
| **Practice Population Deprivation (IMD): % difference vs Q1 (NOT included in Multiple hypothesis test)** |  |  |  |  |  |  |  |  |
| Q2 Income deprivation | 2.0 | 1.8 | 5.2<sup>^</sup> | 4.6 | 2.5 | 4.1<sup>^</sup> | 0.6 | 1.2 |
| Q3 Income deprivation | 1.7 | 3.5 | 9.1<sup>^</sup> | 6.5<sup>^</sup> | 1.5 | 2.8 | 6.0 | 6.4 |
| Q4 Income deprivation | 1.6 | 4.3 | 5.2 | 3.8 | 0.2 | 1.9 | 7.5 | 8.7 |
| Q5 Income deprivation (most deprived) | 1.4 | 5.5 | 9.6<sup>^</sup> | 7.0 | -1.2 | -0.1 | 12.9<sup>^</sup> | 13.7<sup>^</sup> |
| **% change in outcome per 1 SD increase in each predictor, relative to outcome baseline means** |  |  |  |  |  |  |  |  |
| **Practice Characteristics: QOF & Access (Multiple hypothesis - Šidák corrected)** |  |  |  |  |  |  |  |  |
| QOF points | 0.3 | 0.3 | 0.7<sup>^^</sup> | 0.7<sup>^^</sup> | 0.4 | 0.4 | 0.7 | 0.8 |
| Satisfied in making appointment (%) | -1.3<sup>^^^</sup> | -1.4<sup>^^^</sup> | 1.0<sup>^^</sup> | 1.0<sup>^^</sup> | 0.4 | 0.4 | -1.3<sup>^^</sup> | -1.3<sup>^^</sup> |
| Satisfied with phone access (%) | -0.8<sup>^^</sup> | -0.8<sup>^^</sup> | 0.3 | 0.3 | 0.4 | 0.4 | -0.1 | 0.1 |
| **GP Staffing (Multiple hypothesis - Šidák corrected)** |  |  |  |  |  |  |  |  |
| Proportion Female GP FTE (%) | 1.3<sup>^^^</sup> | 1.3<sup>^^^</sup> | -0.8<sup>^^</sup> | -0.8<sup>^^</sup> | 0.5<sup>^^</sup> | 0.5<sup>^^</sup> | 0.2 | 0.2 |
| GP UK medical qualification (%) | 0.2 | 0.3 | 0.1 | 0.1 | 0 | 0.1 | -0.3 | -0.3 |
| GP FTE per 1,000 patients | 0.8<sup>^^^</sup> | 0.7<sup>^^^</sup> | 0.4 | 0.5 | -0.1 | -0.1 | 0 | 0 |
| Average GP age | -3.2<sup>^^^</sup> | -3.5<sup>^^^</sup> | 2.1<sup>^^^</sup> | 2.2<sup>^^^</sup> | -1.1<sup>^^^</sup> | -1.1<sup>^^^</sup> | 0.5 | 0.4 |
| Nurses FTE per 1,000 patients | -0.3 | -0.5 | -0.3 | -0.2 | 0.4 | 0.3 | -0.2 | -0.4 |
| DPC (non-pharma) FTE per 1,000 patients | 0.2 | 0.3 | -0.7 | -0.7 | -0.1 | -0.1 | -0.2 | -0.1 |
| DPC Pharmacy FTE per 1,000 patients | 0.4<sup>^^</sup> | 0.5<sup>^^</sup> | -0.3 | -0.3 | -0.2 | -0.2 | -0.1 | -0.1 |
| **Cancer incidence and population controls (NOT included in Multiple hypothesis test)** |  |  |  |  |  |  |  |  |
| **Cancer Incidence** |  |  |  |  |  |  |  |  |
| Lagged cancer incidence | 1.9<sup>^</sup> | 1.8<sup>^</sup> | -4.5<sup>^</sup> | -4.6<sup>^</sup> | 0.9<sup>^</sup> | 0.9<sup>^</sup> | -1.7<sup>^</sup> | -1.8<sup>^</sup> |
| **Practice Population** |  |  |  |  |  |  |  |  |
| Female patients (% total) | 3.6<sup>^</sup> | 3.5<sup>^</sup> | -4.2<sup>^</sup> | -4.8<sup>^</sup> | 0.2 | 0 | -0.6 | -1.0 |
| Patients aged 15–44 (% total) | -12.7<sup>^</sup> | -11.0<sup>^</sup> | 3.4 | 5.5 | 0.4 | 0.3 | -2.9 | -4.5 |
| Patients aged 45–64 (% total) | -4.7<sup>^</sup> | -3.9<sup>^</sup> | 5.2<sup>^</sup> | 5.8<sup>^</sup> | -1 | -1.2 | 1 | 0.3 |
| Patients aged 65–74 (% total) | -5.3<sup>^</sup> | -3.8<sup>^</sup> | 11.8<sup>^</sup> | 13.1<sup>^</sup> | -0.9 | -0.7 | 7.5<sup>^</sup> | 6.2<sup>^</sup> |
| Patients aged 75–84 (% total) | 15.1<sup>^</sup> | 14.4<sup>^</sup> | 6.2<sup>^</sup> | 6.1<sup>^</sup> | 2.6<sup>^</sup> | 2<sup>^</sup> | 15.1<sup>^</sup> | 16.5<sup>^</sup> |
| Patients aged 85+ (% total) | 2.2<sup>^</sup> | 2.1<sup>^</sup> | 2.9<sup>^</sup> | 3<sup>^</sup> | 0.6 | 0.6 | 10<sup>^</sup> | 9.8<sup>^</sup> |
| Observations & Practices | 39,421 & 6,194 |  | 39,421 & 6,194 |  | 39,404 & 6,194 |  | 39,421 & 6,194 |  |
| R-squared (Model 1 / Model 2) | 0.58 / 0.58 |  | 0.13 / 0.13 |  | 0.04 / 0.04 |  | 0.02 / 0.02 |  |
| F-stat (Model 1 / Model 2) | 569.6 / 169.2 |  | 135 / 40.5 |  | 65.79 / 20.35 |  | 18.74 / 6.65 |  |

Notes: **Model 1** is panel fixed-effects model without interactions; values are OLS coefficients scaled to % change per 1 SD, relative to outcome means. **Model 2** is deprivation-interaction fixed-effects model; values are average marginal effects (AMEs) scaled to % change per 1 SD and averaged over the sample distribution.
We applied the Šidák correction to the 10 staffing and access predictors because these were the modifiable practice factors tested repeatedly in the main, sensitivity, and subgroup analyses (**Supplementary Material Table S.6**). We did not include cancer incidence or patient age and sex composition in this correction, as these variables were used to adjust the models rather than tested as study findings. We also excluded income deprivation because it was the main exposure of interest, not one of the exploratory predictors.
**Supplementary Material Table S.3 and Table S.4** show the full regression results of both models and corresponding confidence interval.
**How to read the percentage:** A value of 2 indicates a change equivalent to 2% of the mean outcome shown at the top of the column. For example, for USC referrals (mean 3,751 per 100,000), a 2% change corresponds to ~75 referrals per 100,000 patients.
<sup>^</sup> = 95% CI excludes zero (nominal significance only). These variables were not tested using Šidák correction because they are largely stable within practice, rather than modifiable by practices. These results are adjusted for the age and sex structure of the practice population.
<sup>^^</sup> = significant at p<0.05 but did not survive Šidák correction, applied across 10 predictors.
<sup>^^^</sup> = significant at p<0.05 after Šidák correction.
The full regression results tables with coefficients and confidence intervals are in **Supplementary Material Section D**.

Income deprivation showed limited association with our cancer diagnostic activity outcomes. Only emergency presentations were consistently higher for practices in the most deprived quintile (12.9–13.7% higher emergency presentation rates, compared to the least deprived quintile).

We also found limited evidence that associations between these predictors and cancer diagnostic outcomes differed across deprivation quintiles. Table 3 and Figure 1 showed that the estimated percentage changes for access and workforce predictors remained largely unchanged after including deprivation interactions in the model. Figure 1 also showed no consistent deprivation gradient in average marginal effects for access and workforce predictors. Average GP age was an exception, as its association with USC referral rates became progressively less negative from Q2 to Q5. Its association with USC PPV peaked at Q3 (+0.5%) and weakened toward both extremes. Its association with USC sensitivity was significant only in the most deprived quintile (-0.2%).

**Figure 1.**
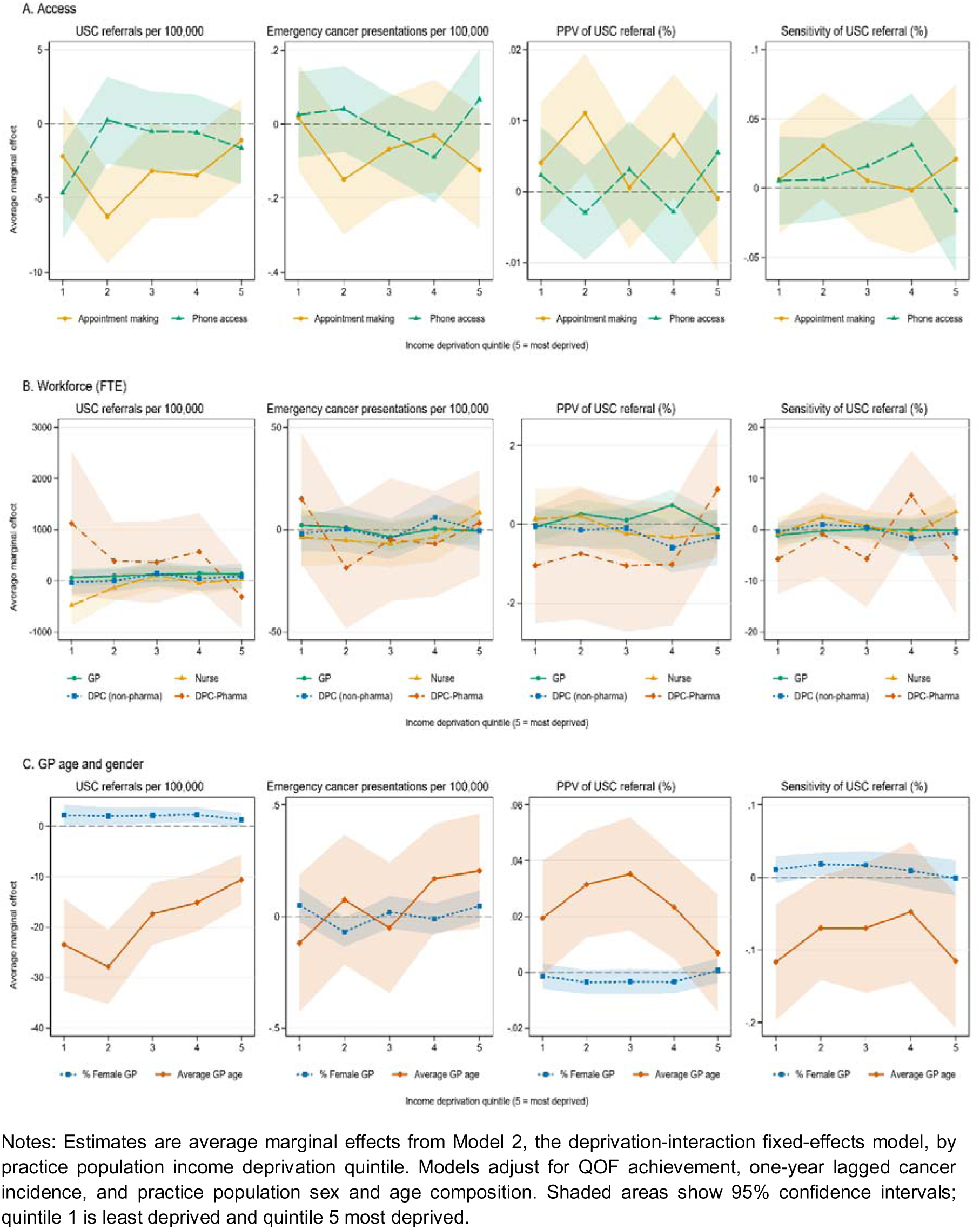
Average marginal effects of access and workforce predictors by income deprivation quintile. Notes: Estimates are average marginal effects from Model 2, the deprivation-interaction fixed-effects model, by practice population income deprivation quintile. Models adjust for QOF achievement, one-year lagged cancer incidence, and practice population sex and age composition. Shaded areas show 95% confidence intervals; quintile 1 is least deprived and quintile 5 most deprived.

Figure 2 shows that the deprivation gap in USC referral rates narrowed over time and had largely closed by 2023, supported by a significant deprivation-by-year joint test using the full 2015–2023 period (F(32,6194)=5.47, p<0.001; Supplementary Material Table S5). Emergency presentation rates remained higher among practices serving more deprived populations, with no clear evidence that this gap changed over time (F(32,6194)=1.17, p=0.238). USC PPV showed no stable deprivation pattern across years (F(32,6194)=2.57, p<0.001), while USC sensitivity varied without a clear deprivation gradient and showed weak evidence of change over time (F(32,6194)=1.37, p=0.078). Compared with the unadjusted trends in Supplementary Material Section C, Figures S1 and S2, adjusted changes in USC referral rates, USC PPV, and USC sensitivity were steeper, suggesting that practice-level characteristics partly masked underlying temporal changes.

**Figure 2.**
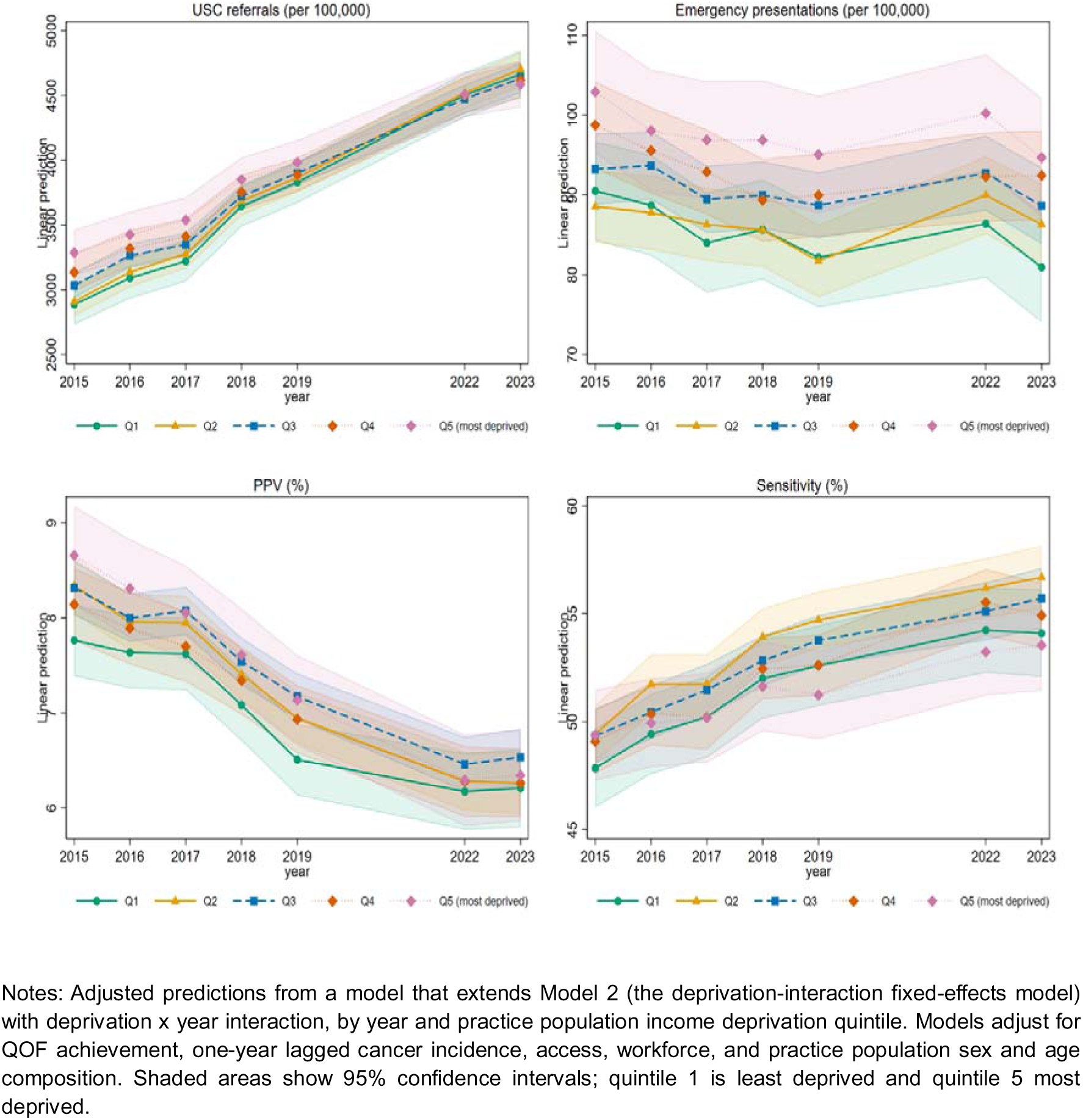
Temporal trends in cancer diagnostic outcomes by deprivation quintile. Notes: Adjusted predictions from a model that extends Model 2 (the deprivation-interaction fixed-effects model) with deprivation x year interaction, by year and practice population income deprivation quintile. Models adjust for QOF achievement, one-year lagged cancer incidence, access, workforce, and practice population sex and age composition. Shaded areas show 95% confidence intervals; quintile 1 is least deprived and quintile 5 most deprived.

### Robustness analyses

The main results were broadly stable across the subgroup and sensitivity checks. Pooling all years available, including 2020–2021, in a single regression moved some coefficients; knowing about the related disruptions, we interpret this as a signal that pandemic years should be treated separately. In the PPV models, adding USC referral volume reduced most access and workforce associations, suggesting that part of these associations may reflect referral volume rather than referral accuracy alone. Deprivation results also changed when year effects were handled differently, in line with the narrowing deprivation gap in USC referrals over time (**Supplementary Materials Table S.7**). Overall, using deprivation deciles instead of quintiles led to the same conclusions, with more nuance for USC PPV. Full results are reported in **Supplementary Material Section E, Figures S3 and S4, and Tables S.6, Table S.7 and Table S.8.**

## Discussion

### Summary

This study explored how within-practice changes in patient access, workforce composition, and practice population income deprivation were associated with cancer diagnostic outcomes. Overall, we found that within practices: (i) better appointment-making experience was associated with fewer USC referrals; (ii) female GP share was associated with more USC referrals, GP FTE showed a small positive association with USC referrals, and older average GP age was associated with fewer USC referrals, higher USC PPV, and lower USC sensitivity; and (iii) increases in the average deprivation of the practice population were associated with higher emergency presentation rates. There was little evidence that deprivation modified associations between access or workforce factors and cancer diagnostic outcomes, except for average GP age. Differences between deprivation quintiles remained similar year-on-year for USC sensitivity, narrowed for USC referral rates, and were inconsistent for USC PPV.

### Comparison with existing literature

Better patient appointment-making experience over time was associated with fewer USC referrals, consistent with prior cross-sectional evidence on access and continuity (3), while our analysis provides within-practice evidence for this association. One possible explanation is that better appointment access may be linked to more consultations for lower-risk or ambiguous symptoms, where GPs may use reassurance, monitoring, or safety-netting rather than urgent referral (14, 15). A cross-sectional study also found that trust and confidence in the doctor was associated with higher USC sensitivity, while ease of seeing a preferred doctor was associated with lower USC sensitivity (16). In contrast, we found association between appointment-making experience and referral volume but not USC PPV or USC sensitivity.

Higher GP FTE and a higher female GP share were associated with more USC referrals within practices. Older average GP age was associated with fewer USC referrals, higher USC PPV, and lower USC sensitivity after multiple-testing correction. This is broadly consistent with previous cross-sectional evidence linking younger GP age with higher referral rates, lower PPV, and higher USC sensitivity (2). The same study reported a small association between GP sex mix and referral activity (2). In our fixed-effects analysis, the association with GP sex mix was clearer and remained significant after multiple-testing correction. Average GP age may capture differences in clinical experience, symptom appraisal, and referral thresholds when assessing ambiguous cancer symptoms (14, 15). Its association also differed by deprivation, but our practice-level data cannot speak to whether this reflects differences in case-mix, symptom presentation, comorbidity, or referral thresholds found in previous studies (17, 18).

A cross-sectional study found that practices serving more deprived populations had higher USC referral and emergency presentation rates, with no clear association for PPV or sensitivity (2), and that deprivation was associated with higher proportions of emergency cancer diagnoses at practice level (19). Our deprivation findings were consistent with emergency presentation, where the deprivation gradient persisted over time and matched earlier longitudinal evidence of persistent emergency-presentation inequality (20, 21). In contrast, the deprivation gap in USC referrals narrowed over time and had largely closed by 2023. Consistent with this, deprivation was associated with higher referral rates only when year fixed effects were excluded, similar to cross-sectional studies (2). USC PPV showed no stable deprivation gradient over time. Its association with deprivation remained after adjustment for USC referral volume, suggesting that referral volume alone is unlikely to explain the PPV pattern. USC sensitivity showed no clear deprivation gradient over time.

### Implications for research and/or practice

These findings suggest that cancer diagnostic improvement should not rely on referral volume alone. For practices and organisations monitoring referral activity, access measures may be more informative when considered alongside USC PPV, USC sensitivity, and emergency presentation rates. Workforce planning should distinguish GP capacity and GP composition from broader direct patient care staffing, as GP FTE, female GP share, and average GP age showed clearer associations with USC referral behaviour. Higher USC PPV in practices with older average GP age should be interpreted alongside referral volume and USC sensitivity, rather than as evidence of better diagnostic performance on its own. Persistent deprivation inequalities in emergency presentation rates, despite a narrowing referral gap, suggest that equity initiatives should also consider barriers before referral, including symptom recognition, timely help-seeking, and access to consultations. Future research should link patient, consultation, clinician, and practice-level data to test whether these patterns reflect referral thresholds, case-mix, continuity, or practice organisation.

### Strengths and limitations

Unlike previous research that largely focused on single outcomes, patient-level variation, or cross-sectional designs (1–3), this study used a longitudinal approach, exploring within-practice associations over time. This allows bias introduced by time-invariant confounders between practices. Findings were also generally similar across subgroup and sensitivity analyses.

This study has several limitations. First, the associations reported here cannot be interpreted causally. Second, the analysis was conducted at the practice level and cannot identify mechanisms operating within individual consultations. We could not determine which GPs within a practice were associated with the observed associations, how skill mix was deployed, or how referral thresholds varied between clinicians. Practice-level outcomes may also mask within-practice heterogeneity.

Third, the deprivation measure reflects changes in the deprivation composition of a practice’s registered population rather than true changes in area-level deprivation or patients’ material circumstances. Fourth, access measures were based on patient-reported experience rather than objective waiting times, and changes in the wording of the GP Patient Survey questions may affect their comparability over time. Fifth, workforce measures were recorded at a single annual time point. Finally, the data do not directly observe referral decision-making, patient–GP interactions, or local diagnostic capacity.

## Conclusion

Practice-level access and workforce factors were associated mainly with USC referral behaviour, rather than consistent improvements in referral performance. Higher emergency presentation rates during periods when practices served populations in the most deprived quintile persisted despite a narrowing deprivation gap in USC referral rates. These findings suggest that efforts to improve early cancer diagnosis should not focus on referral volume alone. Monitoring USC referrals alongside USC PPV, USC sensitivity, and emergency presentation rates may help identify where further support is needed for access, workforce capacity, symptom recognition, and timely help-seeking.

## Data Availability

Metadata and the code for data processing and analysis will be available in our repository or available upon request from the corresponding author.

https://github.com/HOPE-UoM/ACED/tree/main

## Ethical approval

This study used publicly available, practice-level aggregate data and therefore did not require ethical approval.

## Acknowledgment

We are grateful to colleagues and participants at the 2026 BJGP Conference, the 2026 Ca-PRI Conference, the 2026 SAPC Annual Scientific Meeting, and the University of Manchester HOPE seminar for helpful comments on earlier versions of this work.

## Data Availability Statement

Software code reflecting data processing and analyses reported in the paper will be made available on an online repository.

