## Supplementary Material for "Inequalities and practice-level predictors of cancer diagnostic activity: longitudinal study in England"

### A. Data timing, predictor selection and variable construction

**Data timing**

The dataset combines variables from several sources, so reporting periods differ across variables.

For Fingertips variables, we code each financial year by its first calendar year. For example, 2015/16 is coded as 2015. This applies to all outcomes, QOF achievement, cancer prevalence and incidence, and screening coverage.

For GP Patient Survey (GPPS)-based access indicators, including ease of phone contact and good experience making an appointment, fieldwork runs from January to March and results are reported under a single calendar or survey year label, such as 2015.

For variables derived from the General Practice Workforce data, we use the September snapshot in each year rather than an annual average. These variables include GP, nurse, and DPC staffing; GP age, sex, and UK medical qualification composition; and patient age and sex composition.

Ethnicity and rural/urban classification are measured using 2021 cross-sectional data (2021 Census; 2021 Rural-Urban Classification) and applied uniformly across all study years.

**Table A.1** reports the variables used in the main and robustness analyses, including their derivation and reporting period.

**Linking data sources**

Where a common practice identifier was available, data sources were linked at practice-year level. This linkage brought together Fingertips cancer diagnostic indicators, QOF achievement, GPPS access measures, GP Workforce variables, and registered patient counts. In the linked analysis dataset, each observation represents one general practice in one study year.

Annual variables were aligned to the study year definitions reported in Table A.1. Fingertips financial-year indicators were coded by the first calendar year of the financial year. GPPS indicators were aligned using their survey-year label. GP Workforce variables were aligned using the September snapshot for the corresponding study year.

Practice-level deprivation, ethnicity, and rurality measures used geographic information linked to practices. For deprivation, the practice-level value was calculated as the patient-weighted mean of the 2019 IMD income-domain score across patients' Lower Layer Super Output Areas (LSOAs) of residence. For ethnicity and rurality, LSOA-level 2021 Census and 2021 Rural-Urban Classification data were linked to practices through the LSOAs associated with their registered populations. These cross-sectional measures were then applied to all study years.

In calculating practice-level deprivation, IMD weights were renormalised after excluding LSOAs that could not be matched to an IMD income-domain score. All deprivation quintiles and reported analyses use the corrected, renormalised weights.

After linkage, the analysis dataset was treated as a practice-by-year panel. Lagged cancer incidence was created within practice by taking the previous study year's value, as described in Table A.1. We did not impute missing values in the analysis dataset; models were estimated using observations with complete data for the variables included in each specification.

**Predictor rationale**

Among the GPPS-derived indicators available in Fingertips, we selected ease of phone contact and good experience making an appointment as our access measures. These indicators correspond to two stages at which patients may be delayed before seeing a GP: making initial contact and securing an appointment.

We focus on clinical and direct patient care staffing: GPs, nurses, and DPC staff, including pharmacists. These groups are directly involved in diagnostic decision-making and referral behaviour. Practice list size was not included as a separate covariate because its influence is partly captured through rate-based outcomes, practice population age-composition variables, and the per-1,000-patient standardisation of staffing predictors. We examine practice size separately in subgroup analysis by list-size quartile.

**Data cleaning and extreme values**

Outlying staffing and rate-based values were concentrated in small practices, where a fixed count divided by a small patient list can produce rates that vary sharply. The highest USC referral rate was 18,422 per 100,000 in 2023 and came from one small practice, which also had the second-highest rate in the previous year. The highest GP FTE values came from three small practices. Two were branch sites of larger partnerships, where GPs may work across sites while patients are counted at site level. The third was the sole GP practice for the Isles of Scilly, where island isolation may require more staff per registered patient. To limit the influence of outliers, we dropped practice-year observations in the top 1% of the list-size distribution and the bottom 1% of the average-GP-age distribution, where values were unusually low. These exclusions removed individual outlying years rather than whole practices.

**Table S.1. Variable derivations and reporting periods**

| Variable | Derivation | Time basis / reporting period |
|---|---|---|
| ***Outcomes*** | | |
| **USC referral rate** | Fingertips indicator value91882, used without further transformation. For robustness analyses, site-specific USC referral rates are also used for breast, lower gastrointestinal, lung, and skin cancers. | Fingertips: financial year, April–March |
| **PPV / conversion rate** | Fingertips indicator value91845, used without further transformation. | Fingertips: financial year, April–March |
| **Sensitivity / detection rate** | Fingertips indicator value91347, used without further transformation. | Fingertips: financial year, April–March |
| **Emergency presentation** | Fingertips indicator value91356, used without further transformation. | Fingertips: financial year, April–March |
| ***Predictors*** | | |
| **Income deprivation quintile** | Practice-level deprivation is calculated as the patient-weighted mean of the 2019 IMD income domain score across patients' LSOAs of residence. Within each year, practices are then split into five equal-sized groups. Quintile 1 denotes the least deprived practices and quintile 5 the most deprived practices. | IMD score fixed at 2019; patient-registration weights updated annually |
| **Ease of phone access** | Fingertips GPPS indicator value1942, used without further transformation. | GPPS: fieldwork January–March; calendar/survey year label |
| **Good appointment experience** | Fingertips GPPS indicator value349, used without further transformation. | GPPS: fieldwork January–March; calendar/survey year label |
| **GP FTE per 1,000 patients** | Total GP full-time equivalents divided by total registered patients, expressed per 1,000 registered patients. | GP Workforce: September point-in-time snapshot |
| **Nurse FTE per 1,000 patients** | Total nurse full-time equivalents divided by total registered patients, expressed per 1,000 registered patients. | GP Workforce: September point-in-time snapshot |
| **DPC non-pharmacist FTE per 1,000 patients** | DPC non-pharmacist FTE is calculated as total DPC FTE minus DPC pharmacist FTE. This value is divided by total registered patients and expressed per 1,000 registered patients. | GP Workforce: September point-in-time snapshot |
| **DPC pharmacist FTE per 1,000 patients** | DPC pharmacist full-time equivalents divided by total registered patients, expressed per 1,000 registered patients. | GP Workforce: September point-in-time snapshot |
| **% female GP FTE** | Female GP full-time equivalents divided by total GP full-time equivalents, then multiplied by 100. | GP Workforce: September point-in-time snapshot |
| **Average GP age** | Calculated from GP age-band headcounts using the midpoint of each age band. Open-ended bands are assigned 25 years for <30 and 75 years for 70+. The weighted sum of age-band midpoints is divided by total GP headcount. | GP Workforce: September point-in-time snapshot |
| **% GPs UK-qualified** | GPs whose country of qualification is recorded as UK, divided by total GP headcount and multiplied by 100. Country of qualification comes from the GMC Register linked to the GP Workforce dataset. | GP Workforce: September point-in-time snapshot |
| ***Controls*** | | |
| **% female patients** | Female registered patients divided by total registered patients, then multiplied by 100. | GP Workforce: September point-in-time snapshot |
| **% patients per age band** | Male and female patients in each age band are summed, divided by total registered patients, and multiplied by 100. | GP Workforce: September point-in-time snapshot |
| **Practice list size** | Number of registered patients. | Fingertips: financial year, April–March |

| Variable | Derivation | Time basis / reporting period |
|---|---|---|
| **Lagged cancer incidence** | Cancer incidence from the previous financial year for the same practice. The lag is created after declaring the data as a practice-by-year panel. | Fingertips: financial year, April–March; lagged by one financial year |
| **QOF achievement** | Percentage of total achievable QOF points. Fingertips indicator value295, used without further transformation. | Fingertips: financial year, April–March |
| ***Additional variables and sample restrictions used in robustness and sensitivity analyses*** | | |
| **Rural indicator** | Set to 1 if at least 75% of LSOAs linked to the practice are classified as rural; otherwise set to 0. The rural share is the number of rural LSOAs divided by all linked LSOAs, multiplied by 100. | Static: 2021 Rural-Urban Classification, applied to all study years |
| **Ethnicity tertile** | Practices are split into three equal-sized groups based on the proportion of residents recorded as White British in LSOAs linked to each practice, using the 2021 Census. Tertile 1 has the lowest proportion White British and is labelled “highly diverse”; tertile 3 has the highest proportion White British and is labelled “low diversity”. | Static: 2021 Census, applied to all study years |
| **Site-specific USC referral rates** | USC referral rates for breast, lower gastrointestinal, lung, and skin cancers from Fingertips, used without further transformation. These are used as outcomes in cancer-site-specific robustness analyses. | Fingertips: financial year, April–March |
| **Pre-COVID sample restriction** | Sensitivity analysis restricted to study years 2015–2019, before the COVID-19 pandemic. | Study years 2015–2019 only |
| **COVID period indicator** | Set to 1 for study years 2020 and 2021; otherwise set to 0. In the full-sample sensitivity analysis, this indicator is interacted with ease of phone contact, good appointment experience, and lagged crude cancer incidence. | Calendar/study-year indicator |
| **Screening uptake** | Cancer screening uptake from Fingertips, added as an additional control by cancer site where available. | Fingertips: financial year, April–March |
| **Practice size quartile** | Practices are split into four equal-sized groups based on list size. | Fingertips: financial year, April–March |
| **USC referral volume as control** | USC referral volume is added as an additional regressor only in models where PPV is the outcome. In the sensitivity analysis, it is included as a main effect and interacted with deprivation quintile. | Fingertips: financial year, April–March |

**Table S.2. Transition matrix of IMD income deprivation quintiles, 2015 to 2023**

| 2015 quintile / 2023 quintile | Q1 | Q2 | Q3 | Q4 | Q5 | Total |
|---|---|---|---|---|---|---|
| **Q1** | 1,179 (91.6%) | 105 (8.2%) | 2 (0.2%) | 1 (0.1%) | 0 | 1,287 |
| **Q2** | 28 (2.3%) | 1,077 (87.5%) | 123 (10.0%) | 3 (0.2%) | 0 | 1,231 |
| **Q3** | 0 | 24 (2.0%) | 1,041 (86.8%) | 132 (11.0%) | 3 (0.2%) | 1,200 |
| **Q4** | 0 | 0 | 23 (2.0%) | 1,014 (88.1%) | 114 (9.9%) | 1,151 |
| **Q5** | 0 | 0 | 0 | 35 (3.2%) | 1,071 (96.8%) | 1,106 |
| **Total** | 1,207 | 1,206 | 1,189 | 1,185 | 1,188 | 5,975 |

Note: Rows show practices' IMD income deprivation quintile in 2015, and columns show their quintile in 2023. Each cell reports the number of practices, with row percentages in parentheses. The analysis includes 5,975 practices with valid quintile data in both years. Overall, 90.1% of practices remained in the same quintile. Most changes among the remaining practices were movements by one quintile.

### B. Fixed Effect Models

We estimate two regression models using a fixed effects within estimator. In the first model, income deprivation is included as a predictor:

$$Y_{it} = \alpha_i + \lambda_t + \beta X_{it} + \delta W_{it} + \theta IMD_{it} + \gamma Z_{it} + \varepsilon_{it} \quad (1)$$

Where the outcome is measured for practice i in year t and includes the urgent suspected cancer referral rate per 100,000 population, PPV, sensitivity, and emergency presentation rate. Practice fixed effects capture time-invariant practice characteristics that are not measured in the data. Year fixed effects ($\lambda_t$ ) capture shocks or changes common to practices in the same year.

The access predictors are ease of phone contact and good experience making appointments. The staffing indicators include the proportion of female GP FTE, average GP age in the practice, and total full-time equivalents per 1,000 patients for GPs, nurses, DPC staff excluding pharmacists, and DPC pharmacists. Income deprivation is measured as the practice population-weighted average of the IMD income domain score for patients' LSOAs of residence and is included as deprivation quintiles rather than as a continuous score. Control variables include lagged crude cancer incidence, QOF achievement, and practice population age and sex composition. The error term captures idiosyncratic variation.

We exclude practice list size from the fixed-effects models because the main outcomes are already measured as rates or percentages, and staffing variables are expressed per 1,000 registered patients. Practice fixed effects also absorb stable differences in size between practices. We assess size separately in subgroup analyses by list-size quartile.

In the second model, practice-level income deprivation is interacted with all other predictors and control variables. This allows the estimated associations to differ across deprivation quintiles. The model is:

$$\begin{aligned} Y_{it} = {} & \alpha_i + \lambda_t + \beta_1 X_{it} + \delta_1 W_{it} + \gamma_1 Z_{it} + \theta IMD_{it} \\ & + \beta_2 X_{it}\, x\; IMD_{it} + \delta_2 W_{it}\, x\; IMD_{it} + \gamma_2 Z_{it} x\; IMD_{it} + \varepsilon_{it} \end{aligned} \quad (2)$$

Results are reported as average marginal effects, either overall or by deprivation level. The fixed-effects estimator removes time-invariant differences between practices, whether observed or unobserved. The estimates therefore use within-practice changes over time in outcomes and predictors, including changes in the deprivation composition of the registered population, after adjustment for the control variables. Inference is based on standard errors clustered at practice level.

Equation (2) assumes that year effects are the same across deprivation quintiles. We tested this assumption by interacting year effects with deprivation quintiles and using a joint significance test of the interaction terms, as shown in Equation (3).

$$\begin{aligned} Y_{it} = {} & \alpha_i + \lambda_t + \lambda_t\, x\; IMD_{it} + \beta_1 X_{it} + \delta_1 W_{it} + \gamma_1 Z_{it} + \theta IMD_{it} \\ & + \beta_2 X_{it}\, x\; IMD_{it} + \delta_2 W_{it}\, x\; IMD_{it} + \gamma_2 Z_{it} x\; IMD_{it} + \varepsilon_{it} \end{aligned} \quad (3)$$

We use Equation (3) to examine whether yearly changes differ by deprivation quintile.

### C. Time Trend of Outcomes and Predictors

This section presents descriptive unadjusted time trends for the main outcomes and predictors. Figure S1 shows outcome trends overall and by deprivation quintile. Figure S2 shows predictor trends by deprivation quintile. These plots are intended to show the pattern of change in the data before the regression results are reported.

*Figure S1: Time trends of cancer diagnostic outcomes*

Panel a. Cancer diagnostic outcomes time trends, 2015-2023

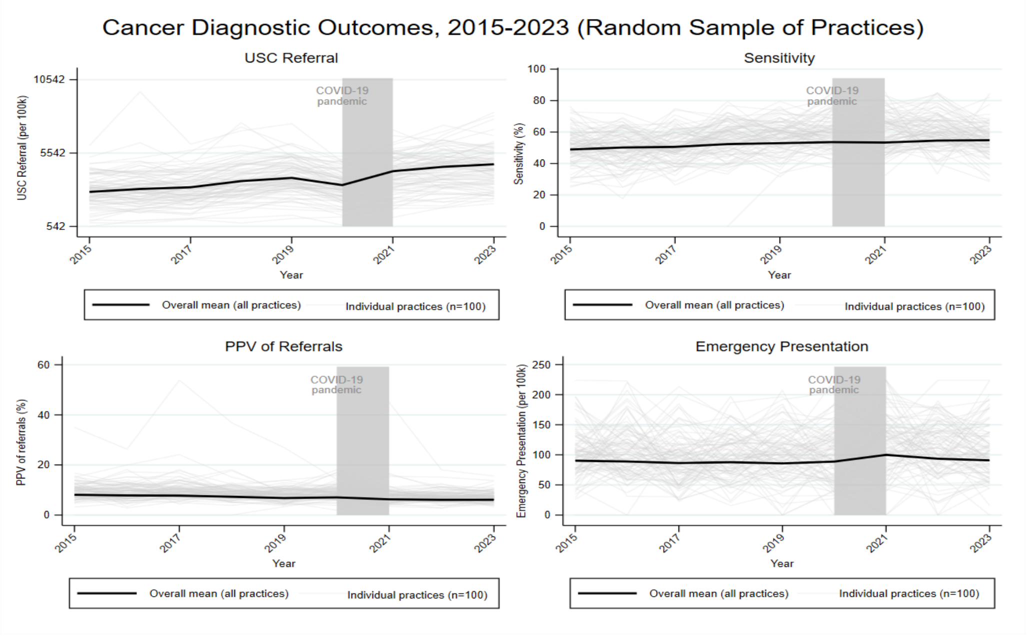


*Note: Thin lines show individual practice trends for a random sample of 100 practices. The thick line shows the overall mean across all practices in each year. The same random seed is used so that the sampled practices can be reproduced.*

Panel b. Cancer diagnostic outcomes by deprivation quintiles

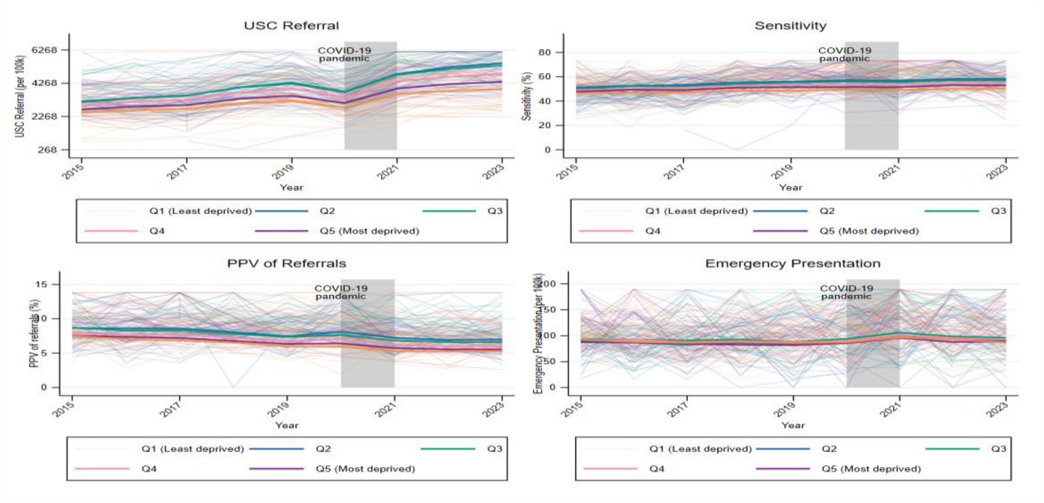


*Note: Thin lines show sampled practice trends, with practices sampled separately within each deprivation quintile (approximately 30 practices per quintile). Thick lines show the mean outcome for all practices in each deprivation*

quintile and year. The sampling is used only for visualisation; quintile means are calculated using all available practices.

#### *Figure S2: Time trends of predictors (workforce and access) by deprivation quintiles.*

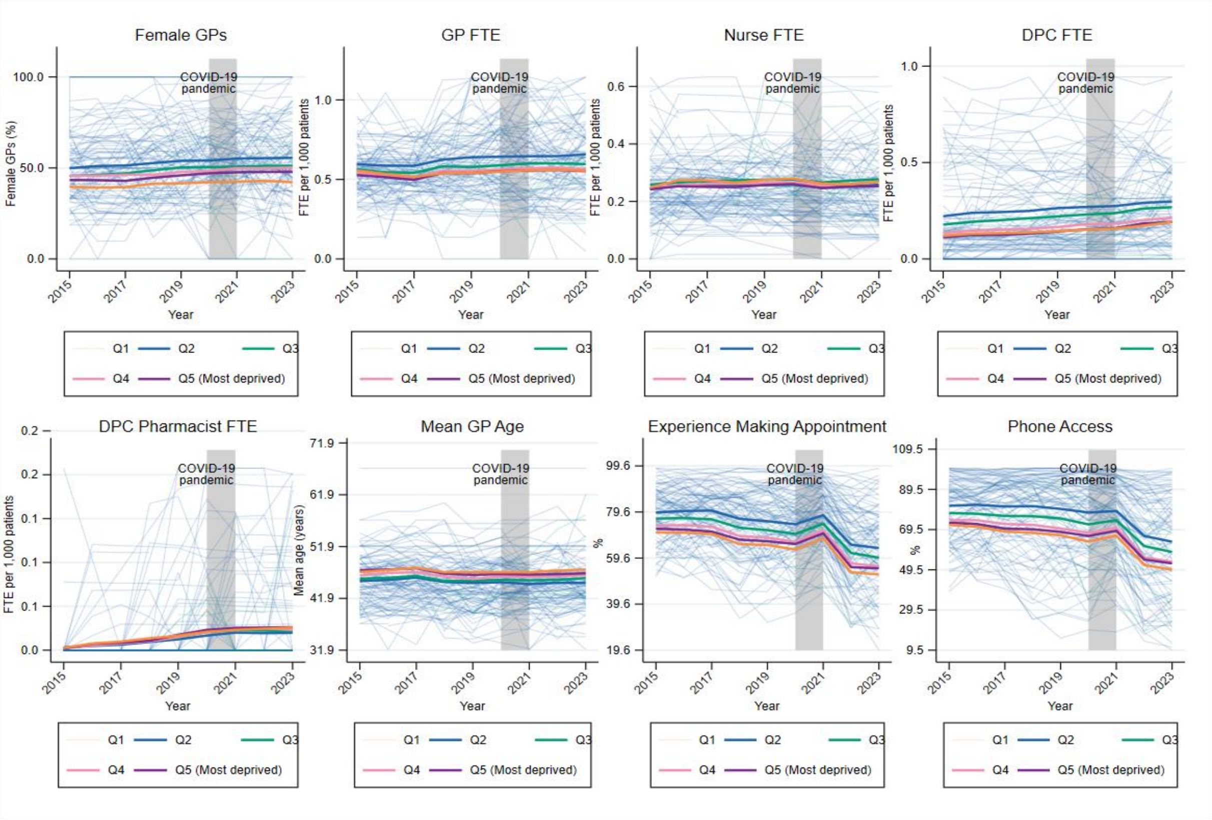


*Note: Thin lines show sampled practice trends, with practices sampled separately within each deprivation quintile (approximately 20 practices per quintile). Thick lines show the mean predictor value for all practices in each deprivation quintile and year. The sampled practices are shown to illustrate within-quintile variation; the thick lines summarise the full analytic sample.*

### D. Regression tables

This section reports the full regression results for the main analyses and the robustness and sensitivity checks. The tables show coefficient estimates, confidence intervals, model fit statistics, and the number of practices included in each specification.

#### Main analysis

##### Model 1

**Table S.3. Model 1 (main effects, no deprivation interaction): full regression results**

| Variable | USC Referral | USC PPV | USC Sensitivity | Emergency Presentation |
|---|---|---|---|---|
| IMD quintile 2 (vs Q1) | 73.874 | 0.373** | 1.280* | 0.547 |
| | (-60.190 - 207.939) | (0.072 - 0.674) | (-0.156 - 2.716) | (-4.756 - 5.850) |
| IMD quintile 3 (vs Q1) | 63.039 | 0.656*** | 0.799 | 5.327 |
| | (-124.940 - 251.018) | (0.256 - 1.056) | (-1.080 - 2.678) | (-2.036 - 12.690) |
| IMD quintile 4 (vs Q1) | 59.404 | 0.376 | 0.101 | 6.709 |
| | (-170.509 - 289.318) | (-0.190 - 0.942) | (-2.480 - 2.681) | (-2.335 - 15.753) |
| IMD quintile 5 (vs Q1) | 51.528 | 0.690** | -0.648 | 11.555** |
| | (-208.154 - 311.210) | (0.034 - 1.346) | (-3.790 - 2.494) | (1.000 - 22.110) |
| Lagged cancer incidence | 0.353*** | -0.002*** | 0.002*** | -0.008*** |
| | (0.271 - 0.436) | (-0.002 - -0.001) | (0.001 - 0.004) | (-0.013 - -0.003) |
| Year: 2016 | 201.812*** | -0.283*** | 1.441*** | -1.967*** |
| | (185.421 - 218.204) | (-0.378 - -0.188) | (1.001 - 1.881) | (-3.426 - -0.508) |
| Year: 2017 | 311.780*** | -0.358*** | 1.836*** | -4.912*** |
| | (291.243 - 332.316) | (-0.455 - -0.260) | (1.377 - 2.296) | (-6.346 - -3.479) |
| Year: 2018 | 681.839*** | -0.845*** | 3.621*** | -5.273*** |
| | (657.721 - 705.956) | (-0.941 - -0.750) | (3.175 - 4.066) | (-6.781 - -3.764) |
| Year: 2019 | 837.857*** | -1.312*** | 4.049*** | -7.259*** |
| | (811.400 - 864.314) | (-1.412 - -1.212) | (3.575 - 4.523) | (-8.781 - -5.737) |
| Year: 2022 | 1,429.435*** | -1.954*** | 5.827*** | -2.344** |
| | (1,385.086 - 1,473.784) | (-2.091 - -1.817) | (5.237 - 6.417) | (-4.333 - -0.354) |
| Year: 2023 | 1,564.008*** | -1.934*** | 5.956*** | -6.055*** |
| | (1,515.521 - 1,612.494) | (-2.077 - -1.790) | (5.331 - 6.582) | (-8.133 - -3.977) |
| QOF achievement | 2.019* | 0.008** | 0.032** | 0.107** |
| | (-0.254 - 4.292) | (0.002 - 0.014) | (0.004 - 0.060) | (0.006 - 0.208) |
| Good appointment experience | -3.100*** | 0.004** | 0.012 | -0.070** |
| | (-4.523 - -1.677) | (0.000 - 0.009) | (-0.009 - 0.032) | (-0.140 - -0.001) |
| Ease of phone access | -1.469** | 0.001 | 0.009 | -0.003 |
| | (-2.799 - -0.139) | (-0.002 - 0.004) | (-0.007 - 0.026) | (-0.060 - 0.053) |
| % female patients | 62.459*** | -0.138*** | 0.036 | -0.237 |
| | (42.284 - 82.634) | (-0.197 - -0.078) | (-0.279 - 0.352) | (-0.992 - 0.517) |
| % patients aged 15–44 | -51.780*** | 0.027 | 0.023 | -0.278 |
| | (-68.367 - -35.194) | (-0.021 - 0.075) | (-0.208 - 0.254) | (-0.870 - 0.313) |
| % patients aged 45–64 | -42.739*** | 0.091*** | -0.130 | 0.216 |
| | (-60.584 - -24.895) | (0.042 - 0.140) | (-0.361 - 0.100) | (-0.492 - 0.923) |
| % patients aged 65–74 | -56.249*** | 0.241*** | -0.135 | 1.911*** |

| Variable | USC Referral | USC PPV | USC Sensitivity | Emergency Presentation |
|---|---|---|---|---|
| | (-81.366 - -31.133) | (0.172 - 0.310) | (-0.418 - 0.148) | (0.949 - 2.874) |
| % patients aged 75–84 | 224.792*** | 0.177*** | 0.528*** | 5.328*** |
| | (195.203 - 254.381) | (0.099 - 0.255) | (0.215 - 0.841) | (4.156 - 6.500) |
| % patients aged 85+ | 70.181** | 0.181** | 0.260 | 7.673*** |
| | (16.180 - 124.181) | (0.028 - 0.335) | (-0.298 - 0.819) | (4.489 - 10.857) |
| % female GP FTE | 1.975*** | -0.002** | 0.011** | 0.007 |
| | (1.198 - 2.752) | (-0.004 - -0.000) | (0.001 - 0.020) | (-0.027 - 0.040) |
| GP FTE per 1,000 patients | 113.424*** | 0.113 | -0.289 | 0.032 |
| | (42.717 - 184.132) | (-0.064 - 0.290) | (-1.211 - 0.632) | (-3.111 - 3.175) |
| Nurse FTE per 1,000 patients | -75.830 | -0.123 | 1.305 | -0.994 |
| | (-199.161 - 47.501) | (-0.447 - 0.202) | (-0.340 - 2.950) | (-6.468 - 4.481) |
| DPC (non-pharmacist) FTE per 1,000 patients | 35.771 | -0.217* | -0.243 | -0.741 |
| | (-79.602 - 151.144) | (-0.470 - 0.036) | (-1.532 - 1.047) | (-5.649 - 4.168) |
| DPC pharmacist FTE per 1,000 patients | 417.460** | -0.574 | -2.520 | -2.626 |
| | (3.816 - 831.104) | (-1.306 - 0.158) | (-6.604 - 1.563) | (-15.790 - 10.538) |
| Average GP age | -17.874*** | 0.022*** | -0.084*** | 0.068 |
| | (-20.946 - -14.801) | (0.013 - 0.032) | (-0.125 - -0.043) | (-0.060 - 0.197) |
| % GPs UK-qualified | 0.216 | 0.000 | 0.001 | -0.008 |
| | (-0.475 - 0.907) | (-0.002 - 0.002) | (-0.008 - 0.010) | (-0.037 - 0.021) |
| Constant | 2,656.659*** | 6.241** | 44.317*** | 35.279 |
| | (965.463 - 4,347.855) | (1.385 - 11.097) | (18.627 - 70.008) | (-27.496 - 98.054) |
| Observations | | | | |
| R-squared | 39,421 | 39,421 | 39,404 | 39,421 |
| Within R² | 0.577 | 0.132 | 0.0446 | 0.0185 |
| F-statistic | 569.6 | 135 | 65.79 | 18.74 |
| Number of practices | 6194 | 6194 | 6194 | 6194 |

*Note: Estimates are from two-way fixed-effects models with practice and year fixed effects. Values in parentheses are 95% confidence intervals based on standard errors clustered at practice level. Statistical significance is denoted by *** p<0.01, ** p<0.05, and * p<0.10. IMD quintile coefficients are not Sidak-corrected. Other modifiable-predictor coefficients are interpreted using the Sidak correction described in the Methods.*

#### Model 2

**Table S.4. Model 2 (deprivation-interacted): IMD quintile main effects and interaction terms**

| Term | USC Referral | PPV | Sensitivity | Emergency Presentation |
|---|---|---|---|---|
| IMD quintile 2 (vs Q1) | 1,430.498 | 1.105 | 61.442* | -41.442 |
| | (-3,414.782 - 6,275.778) | (-11.726 - 13.937) | (-4.356 - 127.239) | (-202.082 - 119.198) |
| IMD quintile 3 (vs Q1) | 4,225.054 | -12.140* | 39.633 | -5.005 |
| | (-1,094.866 - 9,544.973) | (-26.021 - 1.740) | (-28.567 - 107.833) | (-185.039 - 175.029) |
| IMD quintile 4 (vs Q1) | 1,293.287 | -4.578 | 42.878 | -54.036 |
| | (-3,852.459 - 6,439.032) | (-18.311 - 9.154) | (-34.613 - 120.369) | (-226.099 - 118.027) |
| IMD quintile 5 (vs Q1) | 2,725.809 | 4.286 | -5.280 | -17.907 |
| | (-2,564.484 - 8,016.102) | (-10.434 - 19.006) | (-77.775 - 67.215) | (-203.149 - 167.334) |
| Lagged cancer incidence × IMD Q2 | -0.097 | -0.000 | -0.002 | 0.004 |
| | (-0.354 - 0.161) | (-0.001 - 0.000) | (-0.005 - 0.001) | (-0.009 - 0.017) |
| Lagged cancer incidence × IMD Q3 | -0.199 | -0.001 | -0.001 | -0.002 |

| Term | USC Referral | PPV | Sensitivity | Emergency Presentation |
|---|---|---|---|---|
| | (-0.455 - 0.058) | (-0.001 - 0.000) | (-0.004 - 0.003) | (-0.016 - 0.013) |
| Lagged cancer incidence × IMD Q4 | -0.010 | -0.000 | -0.001 | -0.007 |
| | (-0.265 - 0.244) | (-0.001 - 0.000) | (-0.004 - 0.003) | (-0.021 - 0.008) |
| Lagged cancer incidence × IMD Q5 | -0.165 | -0.000 | 0.001 | -0.008 |
| | (-0.433 - 0.104) | (-0.001 - 0.001) | (-0.003 - 0.005) | (-0.024 - 0.007) |
| QOF achievement × IMD Q2 | -8.009** | 0.001 | -0.128*** | 0.079 |
| | (-14.679 - -1.338) | (-0.015 - 0.017) | (-0.198 - -0.058) | (-0.203 - 0.361) |
| QOF achievement × IMD Q3 | 2.093 | -0.007 | -0.042 | 0.008 |
| | (-4.368 - 8.553) | (-0.025 - 0.010) | (-0.120 - 0.035) | (-0.281 - 0.296) |
| QOF achievement × IMD Q4 | 4.100 | 0.001 | -0.010 | 0.077 |
| | (-2.222 - 10.423) | (-0.018 - 0.020) | (-0.102 - 0.081) | (-0.235 - 0.389) |
| QOF achievement × IMD Q5 | 4.148 | -0.000 | 0.028 | -0.279* |
| | (-2.090 - 10.387) | (-0.019 - 0.019) | (-0.058 - 0.114) | (-0.601 - 0.044) |
| Good appointment experience × IMD Q2 | -4.069* | 0.007 | 0.024 | -0.167 |
| | (-8.659 - 0.521) | (-0.005 - 0.019) | (-0.029 - 0.078) | (-0.371 - 0.036) |
| Good appointment experience × IMD Q3 | -0.985 | -0.004 | -0.001 | -0.085 |
| | (-5.621 - 3.651) | (-0.015 - 0.008) | (-0.058 - 0.056) | (-0.285 - 0.115) |
| Good appointment experience × IMD Q4 | -1.288 | 0.004 | -0.008 | -0.049 |
| | (-5.677 - 3.101) | (-0.008 - 0.016) | (-0.067 - 0.052) | (-0.256 - 0.159) |
| Good appointment experience × IMD Q5 | 1.059 | -0.005 | 0.015 | -0.140 |
| | (-3.333 - 5.451) | (-0.018 - 0.008) | (-0.051 - 0.081) | (-0.354 - 0.073) |
| Ease of phone access × IMD Q2 | 4.907** | -0.005 | 0.001 | 0.015 |
| | (0.726 - 9.087) | (-0.015 - 0.004) | (-0.042 - 0.044) | (-0.145 - 0.176) |
| Ease of phone access × IMD Q3 | 4.140** | 0.001 | 0.011 | -0.052 |
| | (0.072 - 8.207) | (-0.009 - 0.010) | (-0.035 - 0.056) | (-0.210 - 0.106) |
| Ease of phone access × IMD Q4 | 4.071** | -0.005 | 0.026 | -0.114 |
| | (0.095 - 8.046) | (-0.015 - 0.005) | (-0.023 - 0.074) | (-0.281 - 0.052) |
| Ease of phone access × IMD Q5 | 3.012 | 0.003 | -0.022 | 0.042 |
| | (-0.914 - 6.938) | (-0.008 - 0.014) | (-0.076 - 0.033) | (-0.134 - 0.217) |
| % female patients × IMD Q2 | 16.322 | -0.007 | -0.523 | 1.456 |
| | (-40.141 - 72.784) | (-0.171 - 0.157) | (-1.280 - 0.234) | (-0.453 - 3.366) |
| % female patients × IMD Q3 | -26.850 | 0.193** | -0.201 | 1.053 |
| | (-90.653 - 36.954) | (0.012 - 0.374) | (-1.032 - 0.630) | (-1.063 - 3.169) |
| % female patients × IMD Q4 | 8.757 | 0.140 | -0.103 | 1.462 |
| | (-52.484 - 69.998) | (-0.041 - 0.320) | (-1.039 - 0.832) | (-0.536 - 3.460) |
| % female patients × IMD Q5 | -6.966 | 0.071 | 0.132 | 0.554 |
| | (-69.172 - 55.241) | (-0.122 - 0.264) | (-0.895 - 1.159) | (-1.667 - 2.774) |
| % patients aged 15–44 × IMD Q2 | -11.235 | -0.027 | -0.283 | -0.186 |
| | (-46.103 - 23.633) | (-0.117 - 0.063) | (-0.769 - 0.203) | (-1.287 - 0.916) |
| % patients aged 15–44 × IMD Q3 | -38.075* | 0.008 | -0.316 | -0.360 |

| Term | USC Referral | PPV | Sensitivity | Emergency Presentation |
|---|---|---|---|---|
| | (-77.298 - 1.149) | (-0.088 - 0.105) | (-0.832 - 0.200) | (-1.663 - 0.944) |
| % patients aged 15–44 × IMD Q4 | -29.672 | -0.054 | -0.457 | -0.186 |
| | (-68.644 - 9.301) | (-0.153 - 0.045) | (-1.004 - 0.090) | (-1.479 - 1.107) |
| % patients aged 15–44 × IMD Q5 | -41.599* | -0.108* | -0.055 | 0.090 |
| | (-83.743 - 0.544) | (-0.221 - 0.005) | (-0.620 - 0.509) | (-1.344 - 1.524) |
| % patients aged 45–64 × IMD Q2 | -30.602 | -0.008 | -0.387 | -0.968 |
| | (-80.911 - 19.706) | (-0.130 - 0.113) | (-0.937 - 0.162) | (-2.857 - 0.921) |
| % patients aged 45–64 × IMD Q3 | -60.351** | 0.071 | -0.367 | -0.914 |
| | (-113.733 - - 6.969) | (-0.061 - 0.203) | (-0.982 - 0.248) | (-2.868 - 1.040) |
| % patients aged 45–64 × IMD Q4 | -52.262** | -0.032 | -0.651* | -1.246 |
| | (-102.511 - - 2.013) | (-0.153 - 0.089) | (-1.338 - 0.037) | (-3.071 - 0.580) |
| % patients aged 45–64 × IMD Q5 | -73.908*** | -0.084 | 0.206 | -0.202 |
| | (-124.289 - - 23.527) | (-0.213 - 0.044) | (-0.446 - 0.858) | (-2.005 - 1.601) |
| % patients aged 65–74 × IMD Q2 | 6.641 | 0.094 | 0.018 | 0.129 |
| | (-53.313 - 66.595) | (-0.043 - 0.231) | (-0.588 - 0.623) | (-2.050 - 2.309) |
| % patients aged 65–74 × IMD Q3 | -14.500 | 0.187** | -0.429 | -0.181 |
| | (-80.494 - 51.495) | (0.020 - 0.353) | (-1.091 - 0.234) | (-2.746 - 2.383) |
| % patients aged 65–74 × IMD Q4 | 14.667 | 0.144 | 0.057 | 1.632 |
| | (-56.783 - 86.116) | (-0.045 - 0.333) | (-0.681 - 0.794) | (-0.912 - 4.176) |
| % patients aged 65–74 × IMD Q5 | 25.103 | 0.084 | -0.379 | 0.385 |
| | (-48.925 - 99.131) | (-0.116 - 0.283) | (-1.222 - 0.463) | (-2.385 - 3.155) |
| % patients aged 75–84 × IMD Q2 | -9.518 | -0.014 | -0.578** | -2.428* |
| | (-70.507 - 51.472) | (-0.146 - 0.119) | (-1.119 - - 0.037) | (-5.049 - 0.193) |
| % patients aged 75–84 × IMD Q3 | -24.138 | -0.020 | -0.274 | -0.993 |
| | (-90.876 - 42.600) | (-0.177 - 0.137) | (-0.886 - 0.338) | (-3.590 - 1.603) |
| % patients aged 75–84 × IMD Q4 | -25.659 | -0.112 | -0.877** | 2.557* |
| | (-100.695 - 49.376) | (-0.308 - 0.083) | (-1.675 - - 0.079) | (-0.370 - 5.484) |
| % patients aged 75–84 × IMD Q5 | -8.117 | -0.051 | -0.903* | 1.751 |
| | (-90.873 - 74.639) | (-0.276 - 0.174) | (-1.808 - 0.002) | (-1.541 - 5.043) |
| % patients aged 85+ × IMD Q2 | -39.285 | -0.132 | 0.013 | 4.988 |
| | (-174.923 - 96.353) | (-0.432 - 0.168) | (-1.278 - 1.303) | (-2.879 - 12.854) |
| % patients aged 85+ × IMD Q3 | -118.875* | -0.067 | 0.155 | 5.022* |
| | (-260.396 - 22.646) | (-0.417 - 0.282) | (-1.172 - 1.481) | (-0.796 - 10.841) |
| % patients aged 85+ × IMD Q4 | -59.795 | 0.007 | -0.059 | -4.973 |
| | (-206.798 - 87.207) | (-0.444 - 0.458) | (-1.766 - 1.648) | (-11.374 - 1.427) |
| % patients aged 85+ × IMD Q5 | -88.877 | -0.127 | 1.065 | 3.386 |
| | (-248.818 - 71.064) | (-0.688 - 0.434) | (-0.819 - 2.949) | (-3.400 - 10.173) |
| GP FTE per 1,000 patients × IMD Q2 | 33.337 | 0.338 | 0.819 | -1.134 |
| | (-203.852 - 270.526) | (-0.169 - 0.846) | (-1.658 - 3.297) | (-10.691 - 8.423) |
| GP FTE per 1,000 patients × IMD Q3 | 55.216 | 0.180 | 1.241 | -5.801 |

| Term | USC Referral | PPV | Sensitivity | Emergency Presentation |
|---|---|---|---|---|
| | (-169.312 - 279.744) | (-0.334 - 0.695) | (-1.335 - 3.817) | (-15.656 - 4.054) |
| GP FTE per 1,000 patients × IMD Q4 | 79.703 | 0.561** | 1.093 | -1.651 |
| | (-135.895 - 295.301) | (0.033 - 1.089) | (-1.777 - 3.964) | (-11.339 - 8.038) |
| GP FTE per 1,000 patients × IMD Q5 | 65.378 | -0.051 | 1.047 | -2.921 |
| | (-138.559 - 269.314) | (-0.590 - 0.488) | (-1.869 - 3.964) | (-12.861 - 7.020) |
| % female GP FTE × IMD Q2 | -0.187 | -0.002 | 0.008 | -0.122** |
| | (-2.758 - 2.385) | (-0.008 - 0.004) | (-0.016 - 0.032) | (-0.220 - - 0.023) |
| % female GP FTE × IMD Q3 | -0.075 | -0.002 | 0.006 | -0.032 |
| | (-2.634 - 2.483) | (-0.008 - 0.005) | (-0.021 - 0.033) | (-0.138 - 0.074) |
| % female GP FTE × IMD Q4 | 0.090 | -0.002 | -0.002 | -0.062 |
| | (-2.375 - 2.555) | (-0.008 - 0.004) | (-0.031 - 0.028) | (-0.167 - 0.043) |
| % female GP FTE × IMD Q5 | -0.923 | 0.002 | -0.012 | -0.004 |
| | (-3.366 - 1.519) | (-0.004 - 0.008) | (-0.042 - 0.019) | (-0.109 - 0.101) |
| Nurse FTE per 1,000 patients × IMD Q2 | 344.953 | 0.075 | 3.303 | -1.550 |
| | (-119.487 - 809.393) | (-0.984 - 1.133) | (-0.871 - 7.477) | (-19.545 - 16.445) |
| Nurse FTE per 1,000 patients × IMD Q3 | 592.208** | -0.370 | 1.538 | -3.255 |
| | (114.868 - 1,069.547) | (-1.366 - 0.626) | (-2.870 - 5.946) | (-21.459 - 14.949) |
| Nurse FTE per 1,000 patients × IMD Q4 | 438.878* | -0.469 | -0.086 | 0.072 |
| | (-18.301 - 896.057) | (-1.557 - 0.619) | (-4.757 - 4.584) | (-18.546 - 18.691) |
| Nurse FTE per 1,000 patients × IMD Q5 | 511.072** | -0.370 | 4.295* | 11.929 |
| | (79.931 - 942.212) | (-1.355 - 0.614) | (-0.603 - 9.192) | (-5.102 - 28.959) |
| DPC (non-pharmacist) FTE per 1,000 patients × IMD Q2 | 31.008 | -0.112 | 1.338 | 2.157 |
| | (-264.968 - 326.983) | (-0.728 - 0.505) | (-1.297 - 3.973) | (-11.354 - 15.668) |
| DPC (non-pharmacist) FTE per 1,000 patients × IMD Q3 | 174.144 | -0.059 | 0.814 | -2.582 |
| | (-165.987 - 514.275) | (-0.750 - 0.631) | (-1.924 - 3.552) | (-15.439 - 10.275) |
| DPC (non-pharmacist) FTE per 1,000 patients × IMD Q4 | 79.116 | -0.563 | -1.318 | 7.849 |
| | (-259.447 - 417.680) | (-1.361 - 0.235) | (-5.341 - 2.706) | (-6.207 - 21.905) |
| DPC (non-pharmacist) FTE per 1,000 patients × IMD Q5 | 126.079 | -0.287 | -0.272 | 1.293 |
| | (-205.530 - 457.689) | (-1.138 - 0.565) | (-5.044 - 4.499) | (-11.459 - 14.044) |
| DPC pharmacist FTE per 1,000 patients × IMD Q2 | -734.049 | 0.301 | 4.927 | -33.735 |
| | (-2,328.539 - 860.441) | (-1.915 - 2.517) | (-5.701 - 15.554) | (-77.526 - 10.056) |
| DPC pharmacist FTE per 1,000 patients × IMD Q3 | -756.887 | -0.006 | 0.012 | -20.087 |
| | (-2,383.674 - 869.900) | (-2.240 - 2.229) | (-11.618 - 11.642) | (-63.905 - 23.732) |
| DPC pharmacist FTE per 1,000 patients × IMD Q4 | -549.186 | 0.027 | 12.464** | -22.036 |
| | (-2,146.781 - 1,048.409) | (-2.116 - 2.170) | (1.392 - 23.536) | (-63.145 - 19.073) |
| DPC pharmacist FTE per 1,000 patients × IMD Q5 | -1,437.519* | 1.935* | 0.141 | -11.844 |
| | (-2,980.535 - 105.497) | (-0.220 - 4.090) | (-12.723 - 13.006) | (-52.723 - 29.035) |
| Average GP age × IMD Q2 | -4.439 | 0.012 | 0.047 | 0.195 |
| | (-15.870 - 6.991) | (-0.016 - 0.040) | (-0.061 - 0.155) | (-0.224 - 0.615) |
| Average GP age × IMD Q3 | 6.093 | 0.016 | 0.047 | 0.068 |

| Term | USC Referral | PPV | Sensitivity | Emergency Presentation |
|---|---|---|---|---|
| | (-4.903 - 17.090) | (-0.013 - 0.044) | (-0.073 - 0.167) | (-0.353 - 0.489) |
| Average GP age × IMD Q4 | 8.335 | 0.004 | 0.069 | 0.290 |
| | (-2.464 - 19.133) | (-0.024 - 0.032) | (-0.056 - 0.194) | (-0.103 - 0.683) |
| Average GP age × IMD Q5 | 12.879** | -0.013 | 0.001 | 0.324 |
| | (2.475 - 23.283) | (-0.042 - 0.017) | (-0.121 - 0.123) | (-0.076 - 0.724) |
| % GPs UK-qualified × IMD Q2 | -0.021 | -0.004 | -0.027** | 0.032 |
| | (-2.145 - 2.102) | (-0.009 - 0.001) | (-0.048 - - 0.005) | (-0.053 - 0.117) |
| % GPs UK-qualified × IMD Q3 | 0.830 | -0.002 | -0.014 | 0.070 |
| | (-1.282 - 2.943) | (-0.008 - 0.004) | (-0.039 - 0.010) | (-0.020 - 0.161) |
| % GPs UK-qualified × IMD Q4 | -0.602 | 0.003 | -0.012 | 0.089** |
| | (-2.604 - 1.401) | (-0.003 - 0.009) | (-0.037 - 0.014) | (0.005 - 0.173) |
| % GPs UK-qualified × IMD Q5 | 1.020 | -0.004 | -0.028** | 0.057 |
| | (-0.912 - 2.952) | (-0.010 - 0.001) | (-0.055 - - 0.001) | (-0.027 - 0.141) |
| Observations | | | | |
| R-squared | 39,421 | 39,421 | 39,404 | 39,421 |
| Within R² | 0.580 | 0.134 | 0.047 | 0.022 |
| F-statistic | 0.580 | 0.134 | 0.0471 | 0.0217 |
| Number of practices | 169.2 | 40.59 | 20.35 | 6.648 |

*Note: Estimates are from two-way fixed-effects models with practice and year fixed effects. The model interacts income deprivation quintile with all predictors and controls. This table reports the IMD quintile main association and the IMD × predictor or control interaction terms. Main associations for access, workforce, and control variables are reported in Table B.3. IMD quintile 1 is the reference group. Interaction terms show differences relative to IMD quintile 1. Values in parentheses are 95% confidence intervals based on standard errors clustered at practice level. Statistical significance is denoted by *** p<0.01, ** p<0.05, and * p<0.10.*

#### Deprivation trend IMD x year

We tested whether the effect of each year is the same across all deprivation quintiles, whether deprivation quintiles moved together over time (H0) or whether certain years affected some quintiles more than others (H1).

**Table S.5. Testing differences in deprivation × year coefficients**

| Outcome | F-statistic | df | p-value | Hypotheses |
|---|---|---|---|---|
| USC Referral | 5.47 | 32, 6194 | <0.001 | Rejected |
| PPV | 2.57 | 32, 6194 | <0.001 | Rejected |
| Sensitivity | 1.37 | 32, 6194 | 0.078 | Not rejected |
| Emergency Presentation | 1.17 | 32, 6194 | 0.238 | Not rejected |

*Note: F-statistics are based on standard errors clustered at practice level. We tested the full i.q_imdincome#i.year interaction block. Rejection indicates that year-to-year changes differ by deprivation quintile for that outcome. These formal tests are included to support the descriptive temporal trends shown in Figure 2 in the manuscript.*

### E. Robustness analyses

For comparison, we also estimated a one-way fixed-effects model without year effects. This assessed whether the estimated associations were sensitive to the inclusion of year fixed effects.

We conducted subgroup robustness analyses by rurality, ethnicity composition, and cancer site. These analyses used the same specification as the main analyses but were estimated separately within each subgroup. For rurality, practices were classified as rural if 75% or more of the LSOAs linked to the practice were classified as rural under the 2021 ONS Rural-Urban Classification. All other practices were classified as urban. This classification used 2021 data and was applied to all study years. The subgroup analysis assesses whether estimated associations are similar across rural and urban practices.

For ethnicity composition, practices were divided into tertiles based on the proportion of residents recorded as White British in LSOAs linked to each practice, using the 2021 Census. Tertile 1 had the lowest proportion White British and was labelled “highly diverse”; tertile 3 had the highest proportion and was labelled “low diversity”. This classification was applied to all study years. The analysis assesses whether estimated associations are similar across practices serving areas with different ethnic compositions.

USC referral models were also re-estimated separately for the four cancer sites monitored in the Fingertips dataset: breast, lower gastrointestinal, lung, and skin cancers. Associations between access, staffing, deprivation, and USC referral may differ by presenting symptom and referral pathway. This analysis tested whether the USC referral results were consistent across cancer sites or concentrated in one or more sites.

We conducted five sensitivity analyses. First, we restricted the sample to 2015–2019, before the COVID-19 pandemic, to estimate associations before pandemic-related disruption. Second, we used the full sample, including the COVID-affected years 2020–2021, and added a binary indicator for these years. This indicator was interacted with ease of phone contact, good experience making appointments, and lagged crude cancer incidence. We included these interactions because patient contact behaviour and diagnostic activity changed during the pandemic and may have changed the association between these predictors and outcomes. Third, we added screening uptake as an additional control variable, by cancer site where available in Fingertips. Higher screening activity may reduce the number of symptomatic patients entering the urgent referral pathway, independently of the main predictors. This analysis assesses whether the main associations are robust to adjustment for screening uptake. Fourth, we repeated the analysis separately within quartiles of practice list size. Smaller practices may have more variable rate-based outcomes because their denominators are smaller. This analysis assesses whether estimated associations are similar across practices of different sizes. Fifth, for models with PPV as the outcome, we added USC referral volume as an additional covariate, both as a main effect and interacted with deprivation quintile. PPV is calculated as diagnosed cancers divided by USC referrals, so a predictor that reduces referral activity could increase PPV even without improving diagnostic accuracy. This “denominator effect” means that PPV associations may partly reflect referral volume rather than diagnostic performance. This analysis assesses whether PPV associations persist after accounting for USC referral volume. Sixth, We also add sensitivity analysis of using deciles rather than quintiles of practice populations' income deprivation.

#### *Figure S3: Sidak significance of key predictor associations across subgroup and sensitivity analyses (interaction model).*

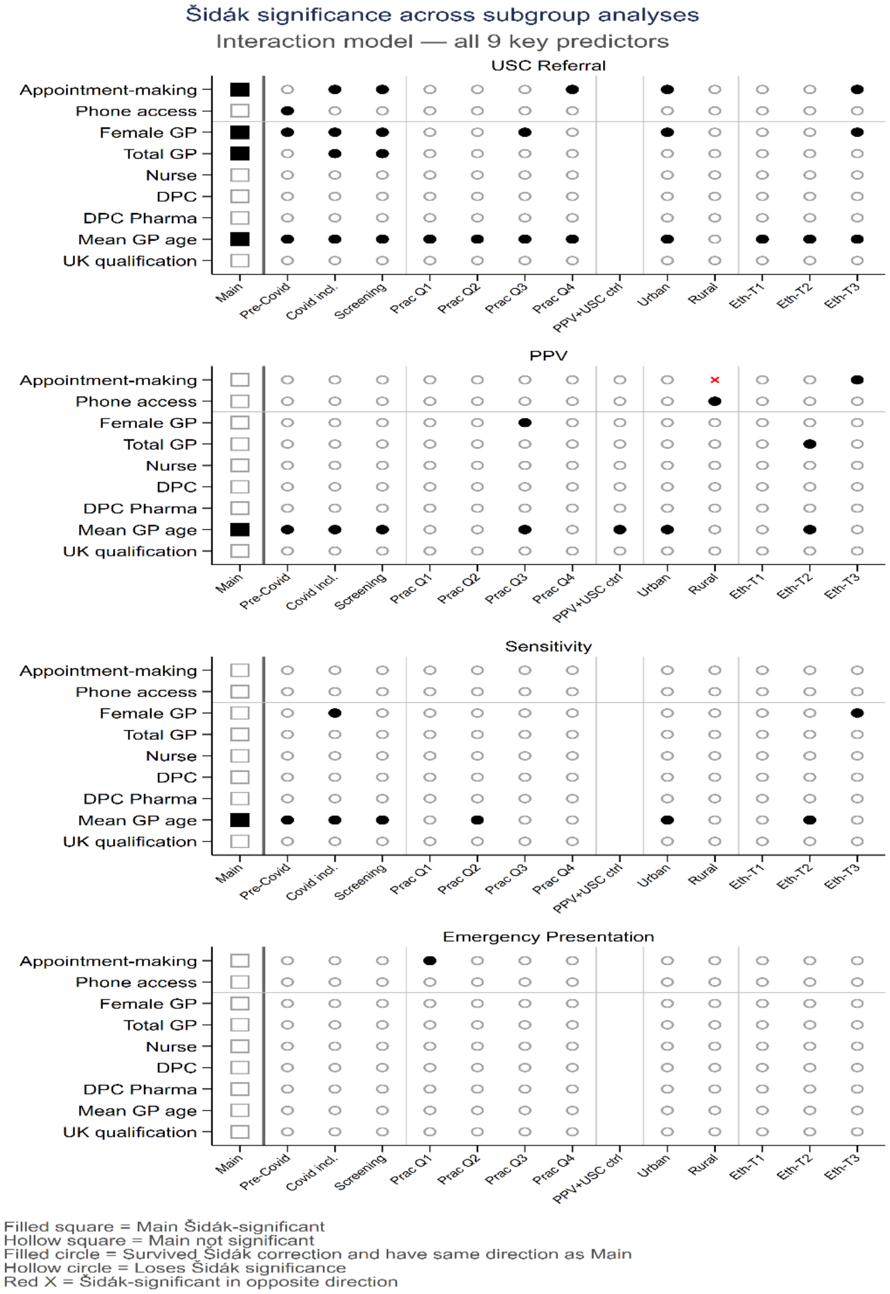


*Note: Filled square = Sidak-significant in the main model. Hollow square = not Sidak-significant in the main model. Filled circle = remained Sidak-significant in the subgroup or sensitivity analysis and had the same direction as the main model. Hollow circle = was not Sidak-significant in the subgroup or sensitivity analysis. Red cross = Sidak-significant in the opposite direction to the main model. Columns are defined as follows: Pre-Covid = 2015–2019 only; Covid incl. = full sample including 2020–2021; Screening = adjusted for screening uptake; Prac Q1–Q4 = practice list-size quartiles; PPV+USC ctrl = PPV models with USC referral volume controlled, PPV outcome only; Urban/Rural = rurality subgroup analyses; Eth-T1–T3 = ethnicity tertiles, where T1 is most diverse and T3 is least diverse. All panels use the fully interacted deprivation-by-predictor specification.*

**Table S.6. Percentage change associated with each predictor (interaction model), by outcome and robustness analysis**

#### USC Referral

| Predictor | Main | Practice FE | Pre-Covid | Covid incl. | Screen-ing | Prac Q1 | Prac Q2 | Prac Q3 | Prac Q4 | PPV+ USC ctrl | Urban | Rural | Eth-T1 | Eth-T2 | Eth-T3 |
|---|---|---|---|---|---|---|---|---|---|---|---|---|---|---|---|
| Appointment-making | -1.4* | -7.6* | -0.6 | -1.3* | -1.5* | -0.8 | -1.1 | -0.8 | -1.5* | n/a | -1.4* | -0.9 | -1.4 | -0.8 | -2.1* |
| Phone access | -0.8 | -3.5* | -1.2* | -0.8 | -0.9 | -3.2 | -1.5 | -0.4 | -0.6 | n/a | -0.9 | +2.3 | -1.6 | -1.1 | +0.1 |
| Female GP | +1.3* | +2.8* | +1.3* | +1.3* | +1.4* | +1.8 | +0.8 | +1.9* | +0.6 | n/a | +1.3* | +1.5 | -0.3 | +1.3 | +2.1* |
| Total GP | +0.7* | +1.9* | -0.1 | +0.8* | +0.8* | +0.5 | +0.4 | +0.9 | +0.9 | n/a | +0.7 | +2.4 | +0.8 | +0.5 | +0.9 |
| Nurse | -0.5 | -1.0* | -0.8 | -0.3 | -0.5 | -0.3 | -0.3 | -0.4 | -0.7 | n/a | -0.5 | -0.1 | -0.9 | -0.6 | -0.1 |
| DPC | +0.3 | +2.5* | +0.5 | +0.4 | +0.2 | +1.3 | +0.2 | -0.1 | +0.2 | n/a | +0.5 | -2.8 | +0.7 | +0.0 | +0.5 |
| DPC Pharma | +0.5 | +2.1* | +0.5 | +0.3 | +0.5 | -0.2 | +0.7 | +0.2 | +0.7 | n/a | +0.4 | +0.4 | +0.7 | +0.0 | +0.7 |
| Mean GP age | -3.5* | -3.5* | -3.2* | -3.4* | -3.5* | -4.8* | -3.4* | -2.8* | -2.0* | n/a | -3.4* | -3.8 | -2.9* | -4.3* | -3.2* |
| UK qualification | +0.3 | +0.1 | +0.2 | +0.2 | +0.3 | -0.3 | -0.1 | +0.7 | +0.0 | n/a | +0.3 | -1.2 | +0.7 | -0.3 | +0.7 |

#### PPV

| Predictor | Main | Practice FE | Pre-Covid | Covid incl. | Screen-ing | Prac Q1 | Prac Q2 | Prac Q3 | Prac Q4 | PPV+ USC ctrl | Urban | Rural | Eth-T1 | Eth-T2 | Eth-T3 |
|---|---|---|---|---|---|---|---|---|---|---|---|---|---|---|---|
| Appointment-making | +1.0 | +5.4* | -0.1 | +1.2 | +1.0 | +1.0 | +0.1 | +1.4 | +0.1 | +1.3 | +1.1 | -11.6* | +0.8 | -0.1 | +2.5* |
| Phone access | +0.3 | +1.6* | +1.5 | +0.4 | +0.3 | +1.4 | +0.6 | +0.5 | +0.8 | -1.2 | +0.3 | +11.4* | +0.8 | +0.2 | -0.4 |
| Female GP | -0.8 | -1.8* | -0.8 | -0.9 | -0.8 | +0.5 | -1.3 | -1.6* | -0.9 | -0.5 | -0.8 | -2.1 | -0.2 | -1.0 | -0.8 |
| Total GP | +0.5 | -0.3 | +0.8 | +0.5 | +0.5 | +0.3 | +0.0 | +0.1 | -0.1 | +0.5 | +0.4 | -0.8 | +0.4 | +1.5* | -0.5 |
| Nurse | -0.2 | +0.2 | -0.1 | -0.1 | -0.2 | -0.9 | +0.6 | -0.0 | -0.7 | -0.2 | -0.2 | -1.8 | +0.5 | +0.1 | -0.7 |
| DPC | -0.7 | -2.2* | -0.6 | -0.8 | -0.5 | -2.8 | -0.4 | -0.1 | +0.2 | -0.5 | -0.6 | -6.7 | +0.8 | -0.3 | -1.4 |
| DPC Pharma | -0.3 | -1.5* | -0.5 | -0.2 | -0.4 | -1.0 | -0.8 | +0.1 | -0.7 | -0.1 | -0.3 | -0.5 | +0.2 | -0.6 | -0.7 |
| Mean GP age | +2.2* | +2.4* | +2.0* | +2.4* | +2.2* | +3.3 | +0.4 | +2.5* | +1.2 | +1.4* | +2.3* | +0.8 | +1.9 | +3.2* | +1.7 |
| UK qualification | +0.1 | +0.3 | +0.3 | +0.0 | -0.1 | +1.6 | -0.5 | -0.8 | +1.1 | -0.2 | +0.1 | +0.1 | +0.0 | +0.3 | -0.4 |

#### Sensitivity

| Predictor | Main | Practice FE | Pre-Covid | Covid incl. | Screen-ing | Prac Q1 | Prac Q2 | Prac Q3 | Prac Q4 | PPV+ USC ctrl | Urban | Rural | Eth-T1 | Eth-T2 | Eth-T3 |
|---|---|---|---|---|---|---|---|---|---|---|---|---|---|---|---|
| Appointment-making | +0.4 | -1.4* | -0.3 | +0.3 | +0.4 | -0.0 | +0.3 | +0.9 | +0.3 | n/a | +0.4 | -2.5 | +0.3 | -0.4 | +1.2 |
| Phone access | +0.3 | -0.1 | +0.6 | +0.4 | +0.4 | +0.8 | -0.1 | +0.5 | -0.2 | n/a | +0.4 | +3.4 | +0.5 | +0.2 | +0.1 |
| Female GP | +0.5 | +1.0* | +0.3 | +0.6* | +0.6 | +0.9 | +0.4 | -0.3 | +0.6 | n/a | +0.5 | +1.0 | +0.4 | -0.0 | +1.1* |
| Total GP | -0.1 | +0.2 | -0.1 | -0.1 | -0.0 | -0.1 | +0.2 | -0.4 | +0.0 | n/a | -0.2 | -0.0 | -0.8 | +0.2 | +0.1 |
| Nurse | +0.3 | +0.1 | -0.0 | +0.2 | +0.2 | +0.4 | +0.3 | +0.7 | -0.2 | n/a | +0.2 | +1.6 | +0.3 | +0.5 | +0.0 |
| DPC | -0.1 | +0.5 | -0.5 | -0.0 | -0.0 | +0.1 | -0.5 | +0.6 | -0.1 | n/a | +0.1 | -5.4 | +1.1 | -0.5 | -0.6 |
| DPC Pharma | -0.2 | +0.3 | -0.2 | -0.1 | -0.2 | -1.3 | +0.5 | -0.2 | -0.4 | n/a | -0.2 | +0.3 | -0.3 | -0.2 | -0.1 |
| Mean GP age | -1.1* | -1.1* | -1.4* | -0.9* | -1.1* | -1.0 | -1.8* | -1.0 | -0.5 | n/a | -1.1* | -1.3 | -1.7 | -1.4* | -0.4 |
| UK qualification | +0.1 | +0.1 | +0.1 | +0.1 | +0.1 | -0.3 | +0.3 | +0.3 | -0.2 | n/a | +0.1 | -0.6 | +0.6 | -0.7 | +0.4 |

#### Emergency Presentation

| Predictor | Main | Practice FE | Pre-Covid | Covid incl. | Screen-ing | Prac Q1 | Prac Q2 | Prac Q3 | Prac Q4 | PPV+ USC ctrl | Urban | Rural | Eth-T1 | Eth-T2 | Eth-T3 |
|---|---|---|---|---|---|---|---|---|---|---|---|---|---|---|---|
| Appointment-making | -1.3 | -0.6 | -1.0 | -1.2 | -1.3 | -6.7* | -0.5 | +0.9 | -0.6 | n/a | -1.0 | -13.0 | -2.9 | -0.4 | -1.2 |
| Phone access | +0.1 | -0.2 | -0.6 | +0.2 | +0.1 | +4.6 | -0.2 | -3.2 | +0.2 | n/a | -0.0 | +6.7 | +1.8 | -0.3 | -0.1 |
| Female GP | +0.2 | -0.1 | +0.5 | -0.3 | +0.1 | +0.6 | +1.0 | +0.9 | -0.9 | n/a | +0.1 | +0.6 | -0.1 | +0.7 | -0.3 |
| Total GP | -0.0 | -0.1 | -0.1 | +0.3 | -0.0 | +1.1 | -1.5 | -1.2 | +0.3 | n/a | +0.1 | -3.2 | +0.8 | +0.2 | -0.4 |
| Nurse | -0.4 | -0.4 | -0.2 | -0.1 | -0.4 | +0.3 | -1.5 | -0.4 | -0.7 | n/a | -0.4 | -2.0 | +0.6 | -1.1 | -0.4 |
| DPC | -0.1 | -0.3 | +0.3 | -0.1 | +0.1 | +1.0 | -0.8 | -0.8 | +0.3 | n/a | -0.2 | +4.8 | +0.9 | +0.7 | -1.2 |
| DPC Pharma | -0.1 | -0.4 | -0.1 | -0.2 | -0.1 | +0.7 | -1.0 | -0.5 | +0.1 | n/a | -0.1 | +1.3 | +0.4 | -0.2 | -0.2 |
| Mean GP age | +0.4 | +0.4 | -0.3 | +0.0 | +0.4 | +0.9 | +0.6 | -0.4 | +0.2 | n/a | +0.4 | -1.9 | +0.1 | +0.9 | +0.3 |
| UK qualification | -0.3 | -0.3 | +0.3 | -0.4 | -0.3 | -0.1 | +0.7 | +0.2 | +0.2 | n/a | -0.2 | -2.2 | -0.6 | +0.5 | -0.4 |

*Note: Values are percentage changes in the outcome per 1 SD increase in the predictor, based on the fully interacted deprivation-by-predictor specification and rounded to 1 decimal place. * indicates that the association is Sidak-significant in that analysis (Sidak-adjusted p<0.05). "n/a" indicates that the analysis does not apply to that outcome; PPV+USC ctrl was run for PPV only. Column definitions match Figure S3: Main = primary two-way fixed-effects model; Practice FE = practice fixed effects only, without year fixed effects; Pre-Covid = 2015–2019 only; Covid incl. = full sample including 2020–2021; Screening = adjusted for screening uptake; Prac Q1–Q4 = practice list-size quartiles; PPV+USC ctrl = PPV models with USC referral volume controlled; Urban/Rural = rurality subgroup analyses; Eth-T1–T3 = ethnicity tertiles, where T1 is most diverse and T3 is least diverse.*

##### Table S.7. Deprivation comparison of practice fixed effects and practice and year fixed effects specifications

| IMD quintile (vs Q1) | USC Referral | | PPV | | Sensitivity | | Emergency Presentation | |
|---|---|---|---|---|---|---|---|---|
| | Practice FE | Practice & Year FE | Practice FE | Practice & Year FE | Practice FE | Practice & Year FE | Practice FE | Practice & Year FE |
| **Q2** | **+7.5%*** | +1.8% | +0.6% | **+4.5%*** | **+5.9%*** | **+4.1%*** | -0.2% | +1.2% |
| **Q3** | **+12.8%*** | +3.4% | +0.03% | **+6.5%*** | **+5.8%*** | +2.8% | +3.5% | +6.4% |
| **Q4** | **+19.4%*** | +4.2% | -6.7% | +3.8% | **+6.5%*** | -1.9% | +5.2% | +8.7% |
| **Q5 (most deprived)** | **+26.0%*** | +5.4% | -7.2% | +7.0% | +5.9% | -0.1% | +9.9% | **+13.7%*** |

*Note: Values are percentage differences relative to Q1, the least deprived quintile, from the non-interaction model. * Indicates that the 95% confidence interval excludes zero. Bold highlights the one-way/two-way contrast.*

#### USC Referral by cancer sites

*Figure S4: USC Referral by cancer sites*

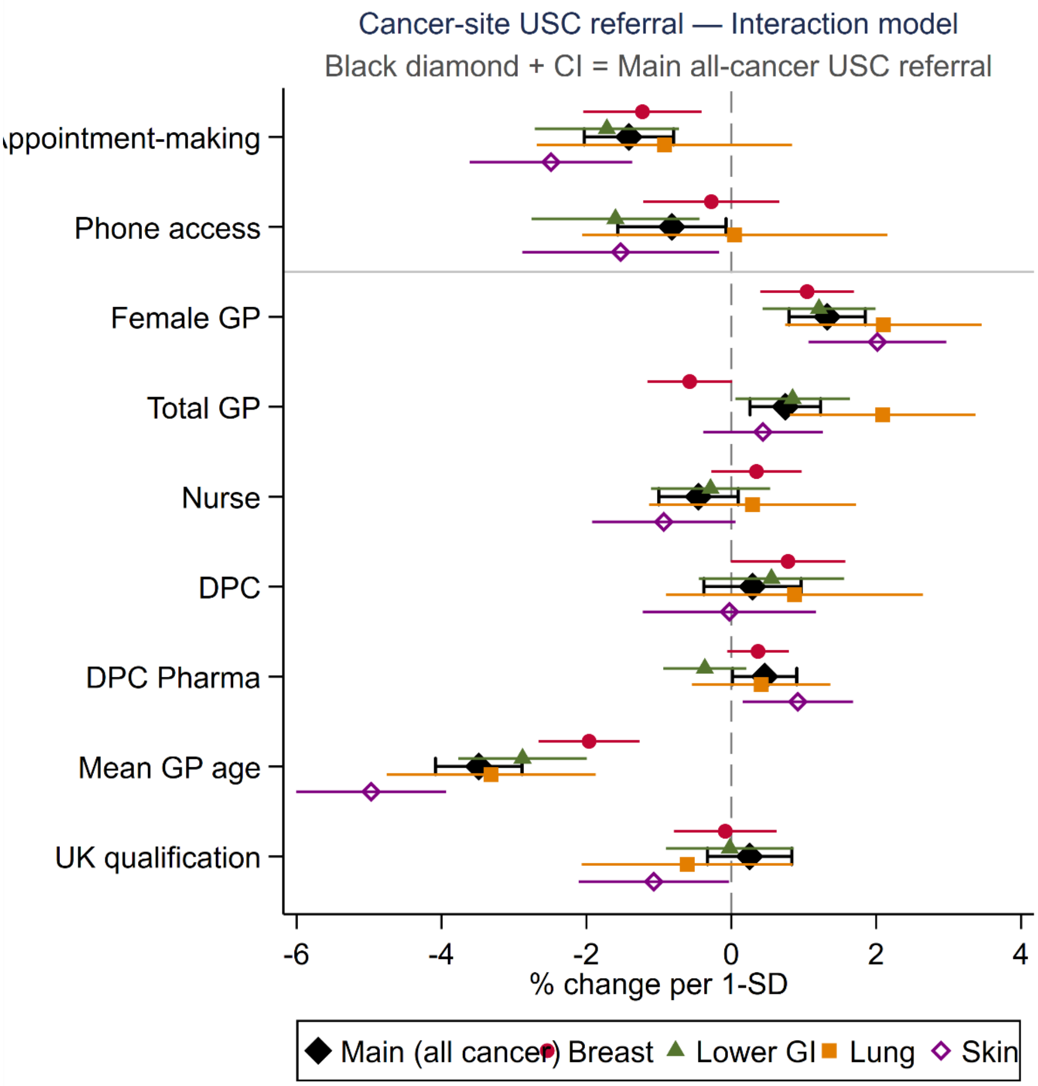


*Notes: The figure presents % change per 1-SD of average marginal effects from cancer-site specific interaction models. Horizontal bars denote 95% confidence intervals. Values are expressed as the percentage change in USC referrals associated with a one standard deviation increase in each predictor. Positive values indicate an increase in referrals and negative values indicate a decrease.*

#### Income deprivation measured in deciles

Because deciles could better capture the shape of the deprivation association, we repeated the analysis with income deprivation split into deciles (D1–D10, D1 least deprived) rather than quintiles, using the same specifications applied to quintiles: practice fixed effects and practice and year fixed effects.

Table E.3 shows that decile analyses gave similar conclusions to quintiles for USC referral, USC sensitivity, and emergency presentation: associations were large and mostly significant under practice fixed effects alone, but attenuated substantially, and were rarely significant, once year fixed effects were added. The main added detail was for USC PPV, where significance under two-way fixed effects was concentrated in D3–D6 and D10, with D7–D9 not reaching significance. Quintiles remain the primary specification for consistency and ease of interpretation.

**Table S.8. Sensitivity of the deprivation effect to fixed-effects specification, decile basis**

| IMD decile (vs D1) | USC Referral | | PPV | | Sensitivity | | Emergency Presentation | |
|---|---|---|---|---|---|---|---|---|
| | Practice FE | Practice & Year FE | Practice FE | Practice & Year FE | Practice FE | Practice & Year FE | Practice FE | Practice & Year FE |
| **D2** | +1.9% | -0.8% | +2.0% | +3.9% | **+3.5%*** | +2.6% | +0.6% | +1.4% |
| **D3** | **+7.7%*** | +1.1% | +4.3% | **+8.9%*** | **+7.2%*** | **+5.1%*** | -0.0% | +2.0% |
| **D4** | **+16.1%*** | +2.7% | +0.9% | **+10.2%*** | **+7.8%*** | +3.6% | -0.2% | +3.4% |
| **D5** | **+19.8%*** | +2.3% | +2.0% | **+14.2%*** | **+8.2%*** | +2.7% | +3.9% | +8.6% |
| **D6** | **+24.9%*** | +2.4% | -2.9% | **+12.8%*** | **+9.9%*** | +2.9% | +2.9% | +8.7% |
| **D7** | **+31.5%*** | +2.3% | -11.4% | +9.0% | **+10.5%*** | +1.7% | +3.2% | +10.0% |
| **D8** | **+38.1%*** | +1.9% | **-17.2%*** | +8.0% | **+10.4%*** | -0.3% | +9.3% | **+17.1%*** |
| **D9** | **+44.2%*** | +1.8% | **-17.1%*** | +12.3% | **+10.7%*** | -1.7% | +14.0% | **+22.4%*** |
| **D10** | **+48.6%*** | -0.8% | **-16.3%*** | **+17.9%*** | **+12.9%*** | -1.5% | +9.9% | **+19.2%*** |

*Note: Values are percentage differences relative to D1, the least deprived decile, from the non-interaction model, rounded to 1 decimal place. * indicates the 95% confidence interval excludes zero (deprivation is not tested for Sidak correction, as it is a structural, non-modifiable predictor).*
